# Frequency and determinants of COVID-19 prevention behaviours: assessment of large-scale programmes in seven countries

**DOI:** 10.1101/2023.11.03.23298032

**Authors:** Sarah Bick, Sian White, Astrid Hasund Thorseth, Max N D Friedrich, Ian Gavin, Om Prasad Gautam, Robert Dreibelbis

**Author notes:** Correspondence to: Robert Dreibelbis, London School of Hygiene & Tropical Medicine, Keppel Street, London WC1E 7HT, United Kingdom.

## Abstract

Pre-existing health and economic challenges mean residents of low- and middle-income countries (LMICs) are likely to be particularly vulnerable to infectious disease pandemics. Limited access to hygiene facilities, water, soap and masks, and dense living environments impeded effective practice of preventive behaviours – handwashing with soap (HWWS), mask wearing and physical distancing – a key line of primary defence against COVID-19. Here we describe a multi-country analysis of prevalence of key hygiene prevention behaviours and their determinants associated with an international non-governmental organisation (WaterAid) hygiene behaviour change programmes for COVID-19 prevention. The goal of this analysis is to inform future outbreak preparedness and pandemic response in LMICs. Cross-sectional household surveys were conducted in October-November 2020 in seven countries where WaterAid worked (Ethiopia, Ghana, Nepal, Nigeria, Rwanda, Tanzania and Zambia). Multivariable mixed-effects regression analyses were used to explore relationships between self-reported behavioural outcomes of interest (handwashing with soap, physical distancing, and mask use) and demographic characteristics, behavioural factors (knowledge, norms, barriers, motives), and exposure to COVID-19 communications. Most respondents (80%) reported increasing their handwashing behaviour after the pandemic, but practice of HWWS at COVID-19-specific prevention moments was low. Mask wearing (58%) and physical distancing (29%) varied substantially between countries. Determinants of key behaviours were identified, including age and socioeconomic status, perceived norms, self-regulation, and the motive of protecting others. These findings highlight that leveraging behaviour-specific emotional drivers and norms, reducing common barriers and promoting targeted messages about specific behaviours and actions individuals can take to reduce risk are necessary to support large-scale behaviour change. Learning from the COVID-19 response to more effectively integrate novel behaviours into existing health promotion will be vital for disease prevention and outbreak resilience.

**Key messages:** *What is already known on this topic:*

- Facilitating COVID-19 prevention behaviours of hand hygiene, mask use and physical distancing in low- and-middle income countries comes with unique challenges
- Identifying effective strategies to promote adoption of key behaviours in diverse contexts over a period of rapid change will be key for future pandemic preparedness

*What this study adds:*

- This multi-country analysis of areas where WaterAid implemented an initial mass media COVID-19 response in 2020 observed lower practice of handwashing at novel COVID-19 prevention moments compared to established moments and variable physical distancing behaviour, and examined behaviour-specific determinants and norms

*How this study might affect research, practice or policy:*

- Renewed focus on identified key drivers of behaviour: targeting critical age-groups and vulnerable populations, increasing descriptive norms and motives of protecting others and respect, and reducing common barriers, with targeted messaging for novel handwashing moments, may be key to ongoing COVID-19 response
- Learning from the rapid COVID-19 response on how well we are able to promote novel behaviours alongside established ones in a variety of contexts can inform future disease prevention and outbreak resilience.

## Introduction

Before mass vaccination, COVID-19 response programmes typically focused on preventive behaviours of hand hygiene, mask use and physical distancing – all seen as a key for reducing transmission and preventing health systems from becoming overburdened. Each of these are different behaviours and facilitating them in low-resource contexts comes with unique challenges. For example, handwashing with soap (HWWS) is a pre-existing, routine behaviour. Global evidence suggests that most people understand the health benefits of HWWS and know how to do it [1]. However, prior to the COVID-19 pandemic, the prevalence of HWWS at critical times (such after using the toilet) in low- and middle-income countries (LMICs) was low [2] and in 28 of the 46 least developed countries only about a quarter of people had access to a basic handwashing facility with soap and water [3]. Water scarcity in many countries also made it hard to prioritise water for handwashing [4–8] and shared water, sanitation and hygiene (WASH) facilities created concerns for COVID-19 transmission [9, 10].

In contrast, mask use was an unfamiliar behaviour to most people prior to the pandemic. Affordable and equitable access to masks (both medical and fabric masks) was limited – particularly during the early stages of the pandemic as medical-grade masks were often prioritised for staff working in health care settings [11, 12]. Hygienic use of masks was challenging in settings where there were high levels of environmental contamination and where laundry is typically done by hand [13]. Physical distancing was also a novel behaviour in most LMICs, often running counter to religious or cultural norms, and it was difficult to enforce or regulate due to large proportions of the population living in densely populated areas and informal settlements [14]. Asking people to reduce unnecessary travel and remain at home came at a much higher socio-economic cost to communities in LMICs due to people, on average, having larger families; smaller houses; being more reliant on daily earnings; having fewer opportunities for collaborating remotely (e.g. access to phones and Wi-Fi); and due to a lack of formal social support mechanisms or financial assistance [14–17].

Communicating about these behaviours or undertaking behaviour change interventions during the pandemic was also particularly challenging in LMICs where health and hygiene promotion programmes have historically prioritised face-to-face interactions with communities due to inequities in access to mass and digital media [18, 19]. Identifying effective strategies to promote adoption of key behaviours in diverse contexts over a period of rapid change was key to improving the ongoing pandemic response and will be key for future pandemic preparedness.

In 2020, WaterAid launched COVID-19 hygiene response programmes in 26 countries. The multi-country approach was underpinned by behavioural theory and a common global strategy but was tailored to national and sub-national contexts. After six months of initial implementation, WaterAid completed a mid-term rapid assessment (MTRA) of targeted COVID-19 behaviours across eight countries, to inform the next phase of the response, of which data from seven were analysed and discussed in this paper. Data were collected about factors that were influencing key COVID-19 prevention behaviours, including socio-demographic factors, exposure to COVID-19 prevention programmes, and other behavioural determinants (e.g. knowledge, norms, barriers, motives), with the aim of informing ongoing pandemic programming and future outbreak resilience. Building off this robust data set, the objectives of the present analysis were to estimate the prevalence of key COVID-19 prevention behaviours – handwashing, mask wearing, and physical distancing – across seven LMIC countries included in the MTRA, and explore relationships with key determinants.

### WaterAid COVID-19 response

WaterAid adapted their existing WASH-related national behaviour change programmes to incorporate COVID-19 specific behaviours. The first phase of the response in May– December 2020 focused on promoting key hygiene behaviours, such as handwashing with soap, covering the mouth and nose when coughing or sneezing, wearing a mask in public places, cleaning / disinfecting frequently touched surfaces and maintaining physical distance. In this first phase, these public health behavioural messages were delivered through non-contact methods such as mass media, digital, social media. The response also included installing handwashing facilities (mostly hands-free, peddle-operated design) and soap in public locations and institutions. Later in the second phase, January–April 2021, communities were reached with face-to-face behaviour change motivational activities including cues, depending on in-country lockdown measures. Further details of the programme delivery and intervention design in each country are found in Supplementary File 1.

## Methods

The MTRA consisted of cross-sectional face-to-face household surveys consisting of closed-ended questions with pre-coded responses in Ethiopia, Ghana, Nepal, Nigeria, Rwanda, Tanzania and Zambia. The data were collected during October and November 2020, with data collection taking up to two weeks to complete in each country. Data were collected by trained field staff in each country; the MTRA survey was completed during the same 4 week period across all 8 countries. Verbal informed consent was collected from each participant at the start of the survey; data collection instruments including consent statements are provided as Supplementary File 2.

### Sampling

In each country, the target population was all adults living within selected geographical areas where WaterAid had implemented its first-phase COVID-19 hygiene promotion and behaviour change response. In each country, the sampling process differed slightly depending on resources, logistical constraints, population data availability and data requirements. Men and women were alternately sampled from one household to the next to ensure an even gender ratio in the sample. Details of the sampling approach are found in Supplementary File 1.

### Measures

Key demographic variables including household and respondent demographics and outcome variables were checked for missing and impossible values. All analyses were conducted by country and at the global level. We used principal component analysis on eight household asset indicators at the country level to construct a household wealth index and divided this into country-specific relative wealth quintiles to use as a covariate in analyses.

Primary outcomes for all analysis were self-reported COVID-19 behaviours targeted by WaterAid’s communications and behaviour change programmes. Specifically, these included handwashing with soap at key moments, mask use, and physical distancing. Descriptions of each outcome and determinant and items used to construct them are available in Supplementary File 3.

Multiple measures of self-reported hand hygiene were collected in the MTRA survey. Questions related to key moments where HWWS was practiced referred to general behaviour with specific recall period (“When do you wash your hands with soap and water?” with multiple responses). Exploratory principle components analysis of self-reported hand hygiene at key moments identified three distinct, related behaviours that were used for future analyses: i) a binary indicator of HWWS after toilet use; ii) a binary indicator of HWWS before eating, and iii) a composite measure of HWWS for prevention of respiratory infection/COVID-19 (COVID-19 HWWS index), scored 0 to 3, consisting of self-reported HWWS after touching frequently-touched surfaces, coming in contact with someone outside the household, or sneezing/coughing. Additionally, we created a binary variable among respondents for self-reported increase in handwashing during the pandemic compared to no change or reduced handwashing.

For mask wearing, we defined an binary indicator based on individuals reporting always wearing a face mask in public spaces vs. reporting sometimes or never wearing a face mask.

For physical distancing, we defined a binary indicator for individuals reporting always practicing physical distancing when in public spaces (one or two meters from others, depending on the country) vs. sometimes or never practicing physical distancing.

A range of possible determinants of self-reported behaviour were captured in the MTRA survey informed by drawing on theoretical frameworks including the RANAS model [20, 21]. These corresponded to broad domains of knowledge, barriers, motives and norms. Questions related to knowledge and barriers were yes/no questions (for example, agreement with “Water is too expensive to purchase for handwashing”) while questions related to motives and norms were 5 point Likert-style questions (for example, with responses ranging from “strongly disagree” to “strongly agree”). To simplify analysis, we grouped all possible questions related to a specific theoretically informed determinants. If data were available on three or more questions, we used Principal Component Analysis (PCA) to create a simple index based on responses. PCA analysis used a tetrachoric correlation matrix for binary variables (knowledge, barriers) and a Pearson correlation matrix for Likert-style responses (motives and norms). Validation involved verification that data presented only one principal component with an Eigenvalue greater than 1, confirmation that similar patterns across countries were observed when performing the analysis at the country level and assessment of internal consistency among items included in indices using Cronbach’s alpha. This was the case for all indices. Indices were rescaled to range from zero to three in order to assess changes in outcomes associated with low, medium and high levels of the determinants. Indicators derived from two items (action knowledge indicators) were a simple total of the two.

Measures of the following determinants were developed during exploratory analysis: *Action knowledge* – knowledge about when to practice a specific behaviour – was operationalised as the total of two dichotomous variables, indicating whether the respondent had named the respective behaviour as a protective behaviour against COVID-19. *Procedural knowledge* referred to participants’ knowledge about how to perform the respective protective behaviour. For HWWS, procedural knowledge referred to knowledge about the correct key situations for HWWS. In line with the three outcomes of HWWS behaviour, we distinguished three dimensions of procedural knowledge: knowing to wash hands before eating, knowing to wash hands after toilet use and knowing to wash hands in situations specifically relevant to prevent a COVID-19 infection. For mask wearing procedural knowledge referred to knowing the situations when to wear a mask. For physical distancing, procedural knowledge referred to knowing its definition, that is staying 1 or 2 meters (depending on the country) from others. Questions on self-regulation – factors that help the individual in managing conflicting goals and distractions when implementing or maintaining a behaviour [20] – were available for handwashing with soap and mask use.

*Barriers* referred to the obstacles that participants reported with regard to the respective protective behaviour. For each target behaviour, participants were asked if anything prevented them from practicing the behaviour, and then asked whether specific barriers were present. Based on exploratory analysis, three types of barriers were distinguished related to handwashing with soap: barriers related to the availability, costs of and access to soap; barriers related to the availability, costs and quality of water; and barriers related to self-regulation such as forgetting or being too busy for HWWS. For mask wearing, barriers included: availability of masks (e.g. costs, lack of knowledge where to buy or how to make a mask); comfort (e.g. difficulties breathing, feeling too hot under a mask); social barriers (i.e. fear of being judged by others); and self-regulation (i.e. forgetting). For physical distancing, barriers included: response efficacy (beliefs as to whether the recommended action step will actually avoid the threat i.e. prevent COVID-19); and barriers related to lack of space.

*Norms* referred to the perceived social pressure to engage in the respective protective behaviour. Two dimensions of norms were distinguished: descriptive norm referred to the respondents’ perception of whether other people engage in the respective protective behaviour (Likert-style responses ranged from “nobody” to “all of them”); and injunctive norm referred to the respondents’ perception of whether other people approved the respondent to engage in the respective protective behaviour (Likert-style responses ranging from “not at all” to “extremely”).

*Motives* described participants feelings and perceived benefits of executing the respective behaviour. For HWWS, the belief that HWWS protects from COVID-19, pride, attractiveness and feeling clean to others were included. For mask wearing, fear of contracting COVID-19 if somebody next to the respondent did not wear a mask, the belief that wearing a mask protects from COVID-19, pride and respect from other were considered. For physical distancing, fear of contracting COVID-19 if not practicing physical distancing, the belief that physical distancing protects from COVID-19, pride and respect were included.

Variables related to self-reported exposure to any COVID-19 communications (not limited to WaterAid communications) were converted to categorical variables for inclusion in analyses, with three levels:

1. no exposure to any COVID-19 communication
2. exposure to COVID-19 communications but not on the behaviour of interest, and
3. exposure to COVID-19 communications on the behaviour of interest.

### Data analysis

We conducted descriptive analysis of primary behavioural outcomes globally, and at the country level. Primary outcomes were disaggregated by gender, age, disability, location, and relative household wealth (Supplementary File 3).

Primary outcomes were assessed with mixed effects regression analyses with the full, multi-country dataset (including country and sampling cluster as random intercepts), with fixed slopes. Poisson regression was used for the outcome of handwashing moments for COVID-19 prevention and logistic regression was used for all other outcomes. We retained four outcomes for the regression analyses due to their relevance for COVID-19 prevention: COVID-19 HWWS index, increase in HWWS behaviour after the COVID-19 pandemic, mask wearing in public spaces, and physical distancing.

Exploratory bivariable regressions were used to explore relationships between each of the potential determinants (demographics, exposure to COVID-19 communications, knowledge and norms related to the targeted behaviour, motives, barriers, household WASH access and exposure/effects of COVID-19) and self-reported behaviours. Among demographic variables, those having a significant association (at the 5% level) with at least one outcome were retained for inclusion in multivariable models – seven demographics were retained (location, gender, age, education, disability status of the respondent, disability status of any member of their household, relative household wealth quintile).

Exploratory multivariable regressions were then used to predict the self-reported behaviours through multiple determinants. Three multivariable regression models were analysed for each outcome: 1) the selected demographics, 2) behavioural factors (motives, barriers, knowledge and norms), and 3) exposure to COVID-19 communications, with models 2 and 3 adjusted for selected demographics. Country-specific workshops were held and insights generated from these results informed subsequent interventions during the COVID-19 hygiene response, including a community-based behaviour change campaign in the second phase.

## Results

### Respondent characteristics

Sampled individuals, villages and geographic location (urban/peri-urban/rural) in the seven countries included are shown in Table 1. In total, 3033 individuals were surveyed across the seven countries. While all respondents lived in urban areas in Ethiopia and Ghana, in other countries respondents resided in a mix of urban, peri-urban and rural areas. Details of individual and household demographics, and household access to water, sanitation and hygiene facilities are in Supplementary File 3.

**Table 1.** Demographic characteristics of respondents and their households by country.

| Country | GLOBAL | Ethiopia | Ghana | Nepal | Nigeria | Rwanda | Tanzania | Zambia |
| --- | --- | --- | --- | --- | --- | --- | --- | --- |
| Individuals | 3033 | 505 | 387 | 497 | 422 | 423 | 395 | 404 |
| Villages / communities | 211 | 8 | 39 | 25 | 48 | 47 | 11 | 33 |
| Geographic area: n (%) |  |  |  |  |  |  |  |  |
| Urban | 1302 (42.9) | 505 (100) | 0 (0) | 144 (29.0) | 183 (43.4) | 85 (20.1) | 154 (39.0) | 231 (57.2) |
| Peri-Urban | 712 (23.5) | 0 (0) | 90 (23.3) | 160 (32.2) | 149 (35.3) | 106 (25.1) | 154 (39.0) | 53 (13.1) |
| Rural | 1019 (33.6) | 0 (0) | 297 (76.7) | 193 (38.8) | 90 (21.3) | 232 (54.9) | 87 (22.0) | 120 (29.7) |

We removed 26 surveys where consent was unclear, and six respondents who self-identified as transgender or did not report their gender (too few observations to include in multivariable analyses).

### Prevalence of COVID-19 preventive behaviours

Figure 1 presents mean values and standard deviations for self-reported HWWS, as well as mask wearing in public spaces, and physical distancing.

**Figure 1.**
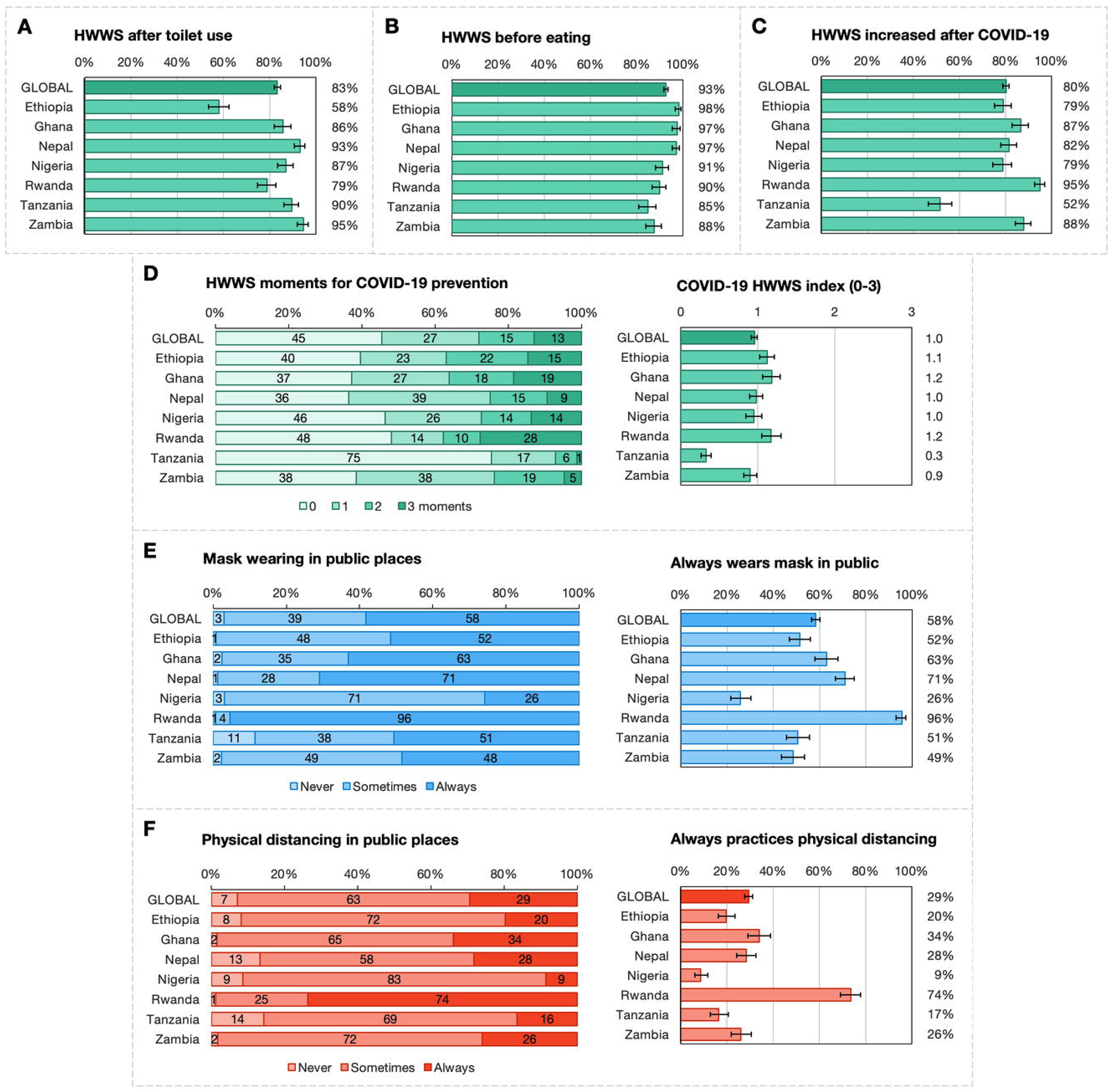
Global and country-level prevalence and means of self-reported COVID-19 prevention outcomes in seven countries where WaterAid worked. A: HWWS after toilet use (%). B: HWWS before eating (%). C: increase in HWWS behaviour after the COVID-19 pandemic (%). D: COVID-19 HWWS index, distribution of key moments and mean values. E: Mask wearing in public spaces (%). F: Physical distancing (%).

More than 80% of participants globally reported that their HWWS practice had increased since the start of the pandemic (Figure 1, panel C). This increase was similar across most countries, except in Rwanda (95%) and Tanzania (52%). Across all countries, more than 80% and more than 90% of participants reported handwashing with soap after toilet use and before eating, respectively (panels A and B). In contrast, in regards to the COVID-19 HWWS index, out of the three possible moments the mean number of moments at which participants reported washing hands with soap was one moment (panel D, right). This finding was similar across countries except for Tanzania with a mean of 0.3 moments. Panel D also presents a more detailed picture of self-reported handwashing behaviour for COVID-19 prevention on the left. Across all data sets, 45% of respondents did not report HWWS at any of the key COVID-19 related junctures. Even fewer respondents practiced any COVID-19 protective HWWS in Tanzania (75% reported no key moments)^a^. 28% of respondents practiced HWWS at two or more key moments for COVID-19. Consistent HWWS in all three situations was reported by few participants across countries ranging from 1% of Tanzanians respondents to 28% of Rwandan respondents.

Prevalence of always wearing a face mask varied considerably by country (panel E, right), with nearly all respondents in Rwanda (96%) always wearing a mask and few (26%) in Nigeria. Globally, only 3% of respondents reported never wearing a face mask (panel E, left); 58% of participants reported always wearing a face mask in public places while 39% reported to sometimes wear a face mask.

While in public, 29% of respondents globally reported to always practice physical distancing (panel F, right). Physical distancing was most practiced in Rwanda, with 74% reporting that they always maintain a distance in public.

### Relationships with demographic characteristics

Table 2 displays multivariable regression analyses of COVID-19 prevention outcomes against selected respondent and household demographics. For the COVID-19 HWWS index, relationship between demographic variables and self-reported behaviour are displayed as incidence rate ratios (IRRs) with corresponding 95% confidence intervals (CIs). For other outcomes, results are displayed as odd ratios OR with corresponding 95% CIs. Gender, age, education, household member with disability and wealth were significantly associated with HWWS moments for COVID-19 prevention. Gender and household wealth were significantly associated with odds of increasing handwashing behaviour after the pandemic.

**Table 2.**
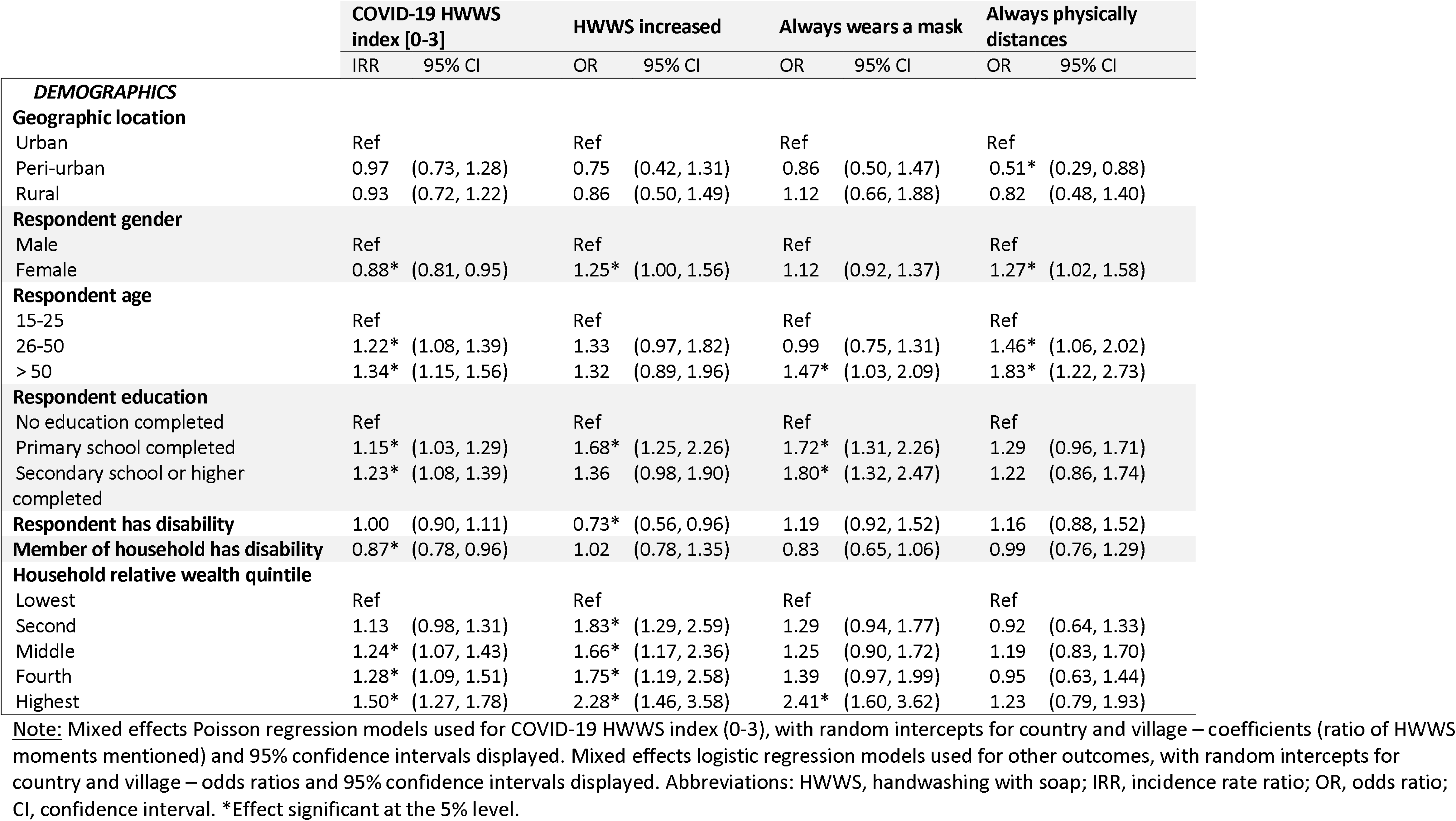
Multivariable regression analyses of COVID-19 prevention outcomes on selected respondent and household demographics in seven countries where WaterAid worked.

Respondents over 50 years old (OR: 1.47) and respondents with at least primary (OR: 1.72) or secondary (OR: 1.80) education were more likely to wear a mask in public (Table 2). Odds of practicing physical distancing was higher among women (OR: 1.27) and increased with age.

### Relationships with behavioural determinants

We conducted multivariate regression analyses in order to quantify how the behavioural factors discussed (knowledge, norms, motives and barriers) related to the behavioural outcomes of interest (Table 3). Procedural knowledge was consistently positively associated with HWWS, and individuals who believed that the behaviour protected others from COVID-19 (action knowledge) tended to report having increased their handwashing behaviour. Fear of COVID-19 was moderately positively associated with an increase in HWWS since the beginning of the pandemic.

**Table 3.** Multivariable regression analyses of COVID-19 prevention outcomes on all behavioural factors (knowledge, barriers, norms and motives), adjusted for key demographics variables, in seven countries where WaterAid worked.

|  | COVID-19 HWWS index [0-3] |  | HWWS increased |  | Always wears a mask |  | Always physically distances |  |
| --- | --- | --- | --- | --- | --- | --- | --- | --- |
|  | IRR | 95% CI | OR | 95% CI | OR | 95% CI | OR | 95% CI |
| **BEHAVIOURAL FACTORS†** |  |  |  |  |  |  |  |  |
| <u>Knowledge</u> |  |  |  |  |  |  |  |  |
| Action knowledge for specific behaviour | 1.02 | (0.89, 1.18) | 1.66* | (1.17, 2.36) | 1.27 | (0.90, 1.79) | 0.95 | (0.67, 1.35) |
| **Procedural knowledge for HWWS** |  |  |  |  |  |  |  |  |
| Moments for prevention of respiratory infection [0–3] | 2.04* | (1.94, 2.14) | 1.10 | (0.93, 1.31) |  |  |  |  |
| After toilet use |  |  | 0.90 | (0.62, 1.30) |  |  |  |  |
| Before eating |  |  | 2.71* | (1.74, 4.23) |  |  |  |  |
| **Procedural knowledge for Mask wearing** |  |  |  |  |  |  |  |  |
| Situations where necessary [0–3] |  |  |  |  | 1.03 | (0.92, 1.15) |  |  |
| **Procedural knowledge for Physical distancing** |  |  |  |  |  |  |  |  |
| Definition of physical distancing |  |  |  |  |  |  | 1.42 | (0.92, 2.19) |
| <u>Barriers</u> |  |  |  |  |  |  |  |  |
| **HWWS** |  |  |  |  |  |  |  |  |
| Soap [0–3] | 0.95 | (0.89, 1.01) | 1.00 | (0.84, 1.18) |  |  |  |  |
| Water [0–3] | 0.98 | (0.91, 1.06) | 0.76* | (0.61, 0.94) |  |  |  |  |
| Self-regulation [0–3] | 0.90* | (0.84, 0.97) | 0.88 | (0.73, 1.06) |  |  |  |  |
| **Mask wearing** |  |  |  |  |  |  |  |  |
| Availability [0–3] |  |  |  |  | 0.69* | (0.59, 0.82) |  |  |
| Comfort [0–3] |  |  |  |  | 0.90 | (0.80, 1.01) |  |  |
| Pride [0–3] |  |  |  |  | 1.11 | (0.69, 1.80) |  |  |
| Self-regulation |  |  |  |  | 0.52* | (0.40, 0.68) |  |  |
| **Physical distancing** |  |  |  |  |  |  |  |  |
| Response efficacy |  |  |  |  |  |  | 0.88 | (0.56, 1.38) |
| Space [0–3] |  |  |  |  |  |  | 0.74* | (0.66, 0.84) |
| <u>Norms</u> |  |  |  |  |  |  |  |  |
| Descriptive norm specific to behavioural outcome [0–3] | 0.99 | (0.93, 1.07) | 1.17 | (0.97, 1.41) | 1.90* | (1.55, 2.33) | 1.78* | (1.47, 2.14) |
| Injunctive norm specific to behavioural outcome [0–3] | 1.02 | (0.94, 1.11) | 1.06 | (0.84, 1.33) | 1.74* | (1.38, 2.19) | 1.42* | (1.11, 1.83) |
| <u>Motives</u> |  |  |  |  |  |  |  |  |
| General fear of COVID-19 | 1.07 | (0.97, 1.17) | 1.33* | (1.06, 1.67) | 1.15 | (0.94, 1.41) | 0.92 | (0.74, 1.14) |
| **Motives specific to behavioural outcome** |  |  |  |  |  |  |  |  |
| Belief that behaviour <u>protects</u> others from COVID-19 [0–3] | 1.01 | (0.90, 1.13) | 1.48* | (1.09, 2.01) | 1.24 | (0.93, 1.66) | 1.04 | (0.73, 1.49) |
| Pride in practicing behaviour [0–3] | 0.99 | (0.87, 1.13) | 1.27 | (0.91, 1.77) | 1.15 | (0.90, 1.46) | 1.11 | (0.81, 1.51) |
| Belief that behaviour makes respondent <u>attractive</u> to others [0–3] | 1.07 | (0.95, 1.20) | 1.22 | (0.90, 1.67) |  |  |  |  |
| Belief that behaviour makes respondent <u>clean</u> to others [0–3] | 1.00 | (0.88, 1.14) | 0.64* | (0.46, 0.89) |  |  |  |  |
| <u>Fear</u> of contracting COVID-19 if behaviour is not practiced [0–3] |  |  |  |  | 1.15 | (0.89, 1.49) | 1.33 | (0.97, 1.83) |
| Respect from community for practicing behaviour [0–3] |  |  |  |  | 1.02 | (0.82, 1.26) | 1.38* | (1.06, 1.80) |

**Note:** Mixed effects Poisson regression models used for COVID-19 HWWS index (0-3), with random intercepts for country and village – coefficients (ratio of HWWS moments mentioned) and 95% confidence intervals displayed. Mixed effects logistic regression models used for other outcomes, with random intercepts for country and village – odds ratios and 95% confidence intervals displayed. Abbreviations: HWWS, handwashing with soap; IRR, incidence rate ratio; OR, odds ratio; CI, confidence interval. \*Effect significant at the 5% level. †Also adjusted for key demographic and household variables (geographic location, respondent gender, age, education level and disability, household member with disability, and relative household wealth quintile)

Normative considerations were associated with mask wearing and physical distancing outcomes: individuals who perceived others to practice the behaviour more frequently (descriptive norm) and who perceived others to approve of practicing the behaviour (injunctive norm) were more likely to report mask wearing and physical distancing.

The effect sizes for descriptive norms were consistently greater than those of injunctive norms for these behaviours. Self-regulation in particular was a significant barrier for HWWS and mask wearing. Respect from the community was positively associated with physical distancing.

### Relationships with COVID-19 communications

Multivariable regression analyses were used to explore relationships between exposure to COVID-19 communications and key behaviours, after adjusting for the selected demographics. Exposure to any communications and exposure to messages specific to each key behaviour were compared to a baseline of no communications heard.

Respondents who recalled hearing messages specific to handwashing practiced 30% more HWWS moments for COVID-19 prevention, and had over three times greater odds of HWWS after toilet use and having increased handwashing behaviour after the pandemic, compared to those who heard no COVID-19 communications (Table 4). The respondent having heard any COVID-19 communications and specific messages on the key behaviour were both associated with significantly higher odds of wearing a mask (ORs: both 2.18). In contrast, recalling general COVID-19 communication was not associated with any of the handwashing outcomes or physical distancing.

**Table 4.** Multivariable regression analyses of COVID-19 prevention outcomes on exposure to general COVID-19 communications, adjusted for key demographics variables, in seven countries where WaterAid worked.

|  | COVID-19 HWWS index [0-3] |  | HWWS increased |  | Always wears mask |  | Always physically distances |  |
| --- | --- | --- | --- | --- | --- | --- | --- | --- |
|  | IRR | 95% CI | OR | 95% CI | OR | 95% CI | OR | 95% CI |
| **TRIGGERS†** |  |  |  |  |  |  |  |  |
| **Exposure to any COVID-19 communications** |  |  |  |  |  |  |  |  |
| No COVID-19 communications heard | Ref |  | Ref |  | Ref |  | Ref |  |
| COVID-19 communications heard but no messages specific to behavioural outcome | 1.04 | (0.74, 1.47) | 1.78 | (0.95, 3.35) | 2.18* | (1.20, 3.96) | 1.32 | (0.64, 2.71) |
| Messages heard specific to behavioural outcome | 1.32* | (1.07, 1.64) | 3.46* | (2.25, 5.34) | 2.18* | (1.33, 3.56) | 1.76 | (0.91, 3.41) |

**Note:** Mixed effects Poisson regression models used for COVID-19 HWWS index (0-3), with random intercepts for country and village – coefficients (ratio of HWWS moments mentioned) and 95% confidence intervals displayed. Mixed effects logistic regression models used for other outcomes, with random intercepts for country and village – odds ratios and 95% confidence intervals displayed. \*Effect significant at the 5% level. †Also adjusted for key demographic and household variables (geographic location, respondent gender, age, education level and disability, household member with disability, and relative household wealth quintile)

## Discussion

In line with other analyses of COVID-19 behaviours in LMICs [22–24], we found that 80% of respondents in the seven studied countries in our analysis reported increasing their handwashing practice in response to the pandemic and over half reported always wearing a mask in public, whereas only 29% practiced physical distancing, a more demanding behaviour that had low levels of adoption throughout the pandemic [24]. The substantial between-country variability in mask wearing and physical distancing behaviours – echoed in other estimates of self-reported mask wearing in LMICs [25–32] ranging from 28% in the Philippines [30] to over 90% in Mozambique [25] – may have reflected differing national pandemic responses, with higher adherence where behaviours were mandatory or already in common practice [12, 33]. Over the study period (October–November 2020), most countries relaxed restrictions to a small degree, with a mean stringency index of 54.4 at the start and 48.4 at the end for the studied countries [34]. We note that countries with the highest mean stringency index (Rwanda; 72.7) had the highest self-reported behaviours, and vice versa for the lowest (Tanzania; 14.8). While handwashing had reportedly increased, a notable finding was the very limited practice of HWWS at key moments associated with COVID-19 prevention compared with established key moments (after toilet use and before eating). Although other analyses of hygiene behaviours during the COVID-19 pandemic often lack detail on key handwashing moments [33], one study in Indonesia also observed a lower frequency of HWWS at similar COVID-19 prevention moments [35].

Regression analyses provided evidence on the demographic characteristics of those who have adopted prevention behaviours and those who might be a focus of future response. Respondent education and relative household wealth were positively associated with multiple behaviours, and residents of peri-urban areas were less likely to practice physical distancing than those in urban areas, perhaps reflecting the spatial and economic limitations to mitigation behaviours in crowded informal settlements and low-resource communities [14, 30]. Additionally, respondents over 50 years old were more likely to practice HWWS for COVID-19 prevention, mask wearing and physical distancing than those under 50. This is corroborated by other research indicating increased reporting of COVID-19 prevention behaviours among older age groups in other LMICs [24, 25, 33, 36, 37]. Young people may have a perceived lower vulnerability to COVID-19, but critically can still transmit pathogens to high risk groups [38]. Other research has indicated that younger people may also be more affected by social norms, and could be effectively motivated by prosocial motives of keeping families and communities safe [38]. In LMICs, which largely have younger populations compared to high-income countries [39], differences in behavioural practice by age and wealth highlight the opportunity to develop targeted and age-appropriate messages for younger populations and more vulnerable segments of the population [12, 39] that utilise evidence of motives and drivers of prevention behaviours.

We explored the various factors that might influence uptake of COVID-19 prevention behaviours. As expected from the associations with wealth and location, we found that mask availability and adequate space were significant barriers to mask wearing and physical distancing behaviours, respectively. Barriers related to self-regulation were significant predictors of HWWS and mask wearing. Future interventions promoting these behaviours might therefore seek to utilise visual cues or ‘nudges’ [40, 41] to provide a reminder to practice behaviours, and increase availability of masks to facilitate behaviours.

Fear of contracting COVID-19 and knowledge of protective behaviours predicted HWWS but no other behaviours, reflecting mixed evidence of the influence of these constructs on prevention behaviours [26, 32, 42–44]. In contrast, norms significantly predicted multiple behaviours, with descriptive norms as consistently stronger predictors than injunctive norms. Uptake of prevention behaviours can be induced by observing similar behaviour in the community or, conversely, discouraged if few are seen to comply [44, 45]. Descriptive norms were strong predictors of prevention behaviours in other settings [31, 35], and an analysis of predictors of COVID-19 behaviours in 28 countries using machine learning identified injunctive norms as the strongest predictors of behavioural adoption, with descriptive norms ranking highly [44]. Public commitments have been widely used to promote HWWS by increasing descriptive norms [46, 47] and should be explored further to promoted HWWS in the context of COVID-19. The motive of protecting vulnerable groups was also highly predictive [44]. These findings point to a focus on increasing perception of others behaviour and targeting behaviour-specific emotional drivers, instead of increasing knowledge, to sustain COVID-19 prevention measures. They also underscore the need for public health interventions centred on communal behaviour and social responsibility in the face of future outbreaks.

We explored the potential influence of COVID-19 communications on self-reported behaviours, and found positive associations between exposure to COVID-19 communications and each of the key behaviours. Exposure to specific messages linking the promoted behaviour directly to COVID-19 prevention or transmission was generally associated with greater effects on self-reported preventive behaviours than those of general COVID-19 communications (not specific to the behaviour). For HWWS, only exposure to specific messages was significantly associated with the outcomes. There is evidence that both specific and non-specific messages can impact behaviour. For example, a COVID-19 intervention in India delivered COVID-19 prevention messages to 25 million recipients and was found to increase adherence to prevention behaviours. This study found that the effects on COVID-19 behaviours that were not directly targeted in the messages increased in the same magnitude to those mentioned [48]. However, messaging may not be sufficient to lead to sustained behavioural change. In a cluster-randomised trial of mask wearing in Bangladesh, a combination of mask distribution, role-modelling and light informal social sanctions were critical to affect behaviour [49]. Efforts to establish social norms around novel behaviours, alongside targeted messages on HWWS moments for COVID-19 prevention, will help stabilise prevention behaviour even as the context in which individuals practice behaviours is rapidly shifting [45].

Non-pharmaceutical interventions such as HWWS, mask wearing and physical distancing are likely to be an important future defence against infectious diseases which have significant outbreak and/or pandemic potential (for example, influenza). Effectively integrating novel behaviours into existing health promotion will be vital for disease prevention and outbreak resilience. For example, hand hygiene plays a critical role in preventing diarrhoeal diseases, trachoma, and respiratory infections [50–52], as well the emergence and spread of other infectious disease outbreaks [53]. However, the typical focus on hand hygiene among caregivers to interrupt faecal-oral transmission of diarrhoeal pathogens will need to be adapted to foster practice of the key moments critical for preventing respiratory viruses we identified as priorities [53]. Innovative research in LMICs during past epidemics, such as testing methods to engage remote populations with novel behaviours during the Ebola crisis, has shaped the current global COVID-19 response [54], and continued learning will enable adaptation of present and future responses.

A strength of the study was the reliance on face-to-face household surveys, which are not subject to the same selection biases as the online and telephone surveys frequently used to assess self-reported behaviour in COVID-19 contexts. We were also able to explore various handwashing moments in the context of a large-scale response programme and across multiple countries. However, there are limitations with using self-reported measures of behaviour, which can often be overreported due to social desirability bias, and reporting of routine behaviours can be particularly affected by recall bias [55]. For example, in a study in Kenya, the 88% prevalence of self-reported mask wearing was reduced to only 10% when observed in practice [56]. Including proxy measures of behaviour may strengthen data collection of this nature in the future. Reporting of behaviour also does not guarantee correct practice. For example, we could not observe if respondents were wearing masks correctly or keeping appropriate distance. The lack of a comparison group or baseline period in the study communities means we cannot make causal links between the intervention and the target behaviours. We used simple ways to aggregate data and indices are a crude representation of the complex psychological and social phenomenon they represent. However, the measures and associations were consistent across countries. We were limited in the number of determinants that could be reflected on in the data. Unfortunately, we were also unable to explore hygiene behaviours among individuals outside of the gender binary due to the limited response rate. Future research should focus on gender-non-conforming individuals and explore how pandemic responses include and address their needs.

## Conclusion

In this multi-country analysis of areas where WaterAid implemented its first-phase COVID-19 response in 2020, we sought to understand prevalence and drivers of self-reported COVID-19 prevention behaviours to improve pandemic learning. We observed high levels of established handwashing moments and mask wearing, but lower practice of handwashing at novel COVID-19 prevention moments and physical distancing, with between-country variation. Our analyses call for a renewed focus on younger and poorer subsections of the population. Pursuing increasing descriptive norms and motives of protecting others and respect, reducing common barriers, with targeted messaging for novel handwashing moments, may help improve and sustain behaviours for reducing the ongoing burden of COVID-19. How well we are able to promote novel behaviours alongside established ones in a variety of contexts may also determine how well we can respond to future emergent pandemic threats.

## Data Availability

The datasets used and analysed during the current study are available from the corresponding author on reasonable request.

## Declarations

### Ethics approval and consent to participate

Approval to collect data has been provided by national or local governments in the WaterAid operating areas. Informed written consent was obtained from all study participants prior to participation. The data were anonymised and full confidentiality has been maintained throughout the process. Ethical approval for analysis of de-identified data was granted by the Ethics Committee of the London School of Hygiene and Tropical Medicine (Reference #22900).

### Contributors

RD and OPG designed and conceptualised the research study. RD, OPG, IG, MNDF and AHT contributed to design and adaptation of data collection tools. WaterAid managed data collection processes. SB, RD, and MNDF analysed the data. SB wrote the first draft of the manuscript. All authors (SB, SW, AHT, MNDF, IG, OPG and RD) reviewed and contributed to subsequent drafts, and read and approved the final manuscript.

### Patient and public involvement

Patients and/or the public were not involved in the design, conduct, reporting or dissemination plans of this research.

### Patient consent for publication

Not applicable.

## Acknowledgements

The WaterAid MTRA Core Group were responsible for conceptualising, overseeing the study design, developing tools, training, feedback on data analysis and on report generation. Special thanks to Core team led by Dr Om Prasad Gautam and supported by Mr Ian Gavin, Ms Lara Kontos, Ms Ellen Greggio, Mr Sabir Hussain, and Mr Ben Robinson.

This report summarises the findings and hard work of the WaterAid county programmes involved. Particular thanks to Bwalya Nachula, Matilda Akua Afriyie, Khakindra Bhandari, Jesse Danku, Lorkumbur Emmanuel, Zinash Kefale, Sunil Koirala, Twaha Mubarak, Rebecca Stanley, Olivier Ndizeye, Brenda Tembo, Raymond Hamoonga and local research partners in each country. The regional staff includes Thérèse Mahon, Tidiane Diallo, Elijah Adera and Ronnie Murungu.

Special thanks to our national and district level government partners who approved and facilitated the study. Thanks to all the respondents for their valuable time and information.

The COVID-19 Hygiene Hub (https://hygienehub.info/en/about/) at LSHTM provided technical support to the questionnaire design (reviewed tools), analysis and multi-country report writing. RANAS conducted country analysis and wrote the country reports.

Findings from this work were presented at the UNC Water and Health conference in October 2021, and Africa Water and Sanitation Week in November 2021.

## Footnotes

a At the time of the survey the government of Tanzania did not recognise COVID-19; therefore, the campaign was framed slightly differently as prevention behaviours for communicable diseases.

