## Supplementary File 1 for "Frequency and determinants of COVID-19 prevention behaviours: assessment of large-scale programmes in seven countries"

### Supplementary File 1: Supplementary notes

#### Note S1: Programme design and delivery

WaterAid’s hygiene response to COVID-19 expected to achieve and contribute the following:

- Improve public awareness and adoption of key hygiene behaviours directly linked to COVID-19 prevention.
- Improve access to handwashing facilities in public places and institutions.
- Contribute to WASH sector coordination for COVID-19 response.
- Contribute to reducing the spread of COVID-19

WaterAid focused on following key hygiene behaviours part of the first phase response (May 2020 to April 2021):

I. **Handwashing with soap:** Frequently washing both hands with soap and water at least 20 seconds. Hand washing should be practiced before cooking and eating; before touching the nose/face; after going to the toilet; after exposure with any dirt/dust/fluids; after coming into contact with frequently touched surfaces; and during, before and after taking care of a sick person.
II. **Respiratory hygiene:** Covering the nose and mouth when coughing and sneezing (sneezing or coughing into the elbow and disposing of the tissue into a bin if it has been used) to be followed by handwashing with soap. Wearing a mask in public.
III. **Physical distancing:** Avoid close contact and maintain two metre (one metre in some countries) distance between yourself and other people. Maintain physical distancing, such as avoiding group gatherings, reducing all non-essential travel and using non-contact greetings.
IV. **Surface cleanliness:** Cleaning and disinfecting frequently touched surfaces regularly, such as door handles, mobile phones and light switches, using disinfectant.
V. **Isolate/referral:** Stay at home if you feel unwell. If you have coronavirus symptoms (high fever, new, continuous cough, difficulty breathing or loss of taste and smell) seek medical attention in advance. Follow your Ministry of Health’s advice.

The programme design used a scaled down version of the Behaviour Centred Design (BCD) approach ^[1]^ – “ABCDE (Assess, Build, Create, Deliver, Evaluate)” – and its methodology to enable identification of the most effective interventions and target the most influential motives for each of the settings in which this programme was implemented. These interventions were underpinned by a behaviour-centred Theory of Change, below.

The interventions focused on changing four key behaviours (I-IV) to help reduce the spread of COVID-19. The hygiene intervention package was designed to change behaviour by: a) changing the **environment** where the behaviour happens (through the placement of behavioural products such as handwashing facilities, visual cues and nudges), b) changing sub-conscious thinking by targeting **motives** and emotions linked to key behaviours (such as affiliation, social status, nurture and disgust with coronavirus), c) and changing **social norms** linked with specific behaviours for habit formation.

Theory of change for the WaterAid hygiene response to COVID-19:
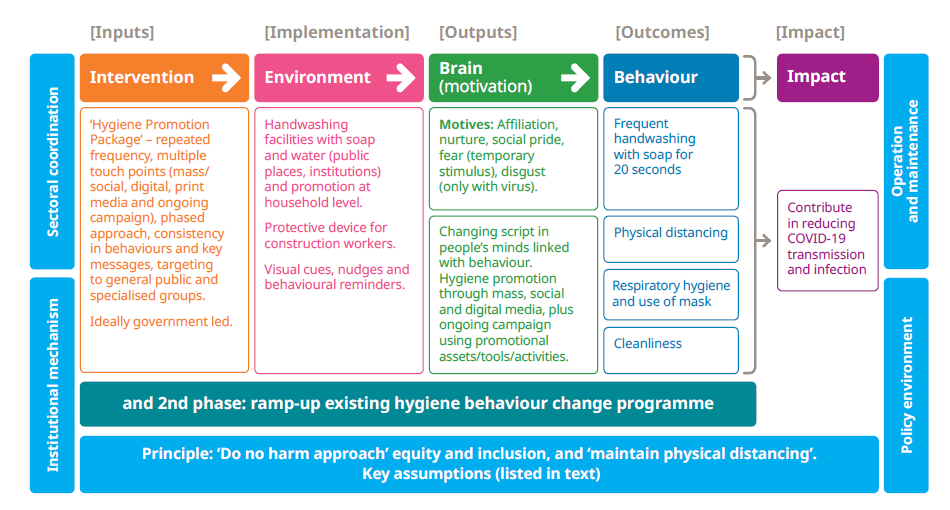

WaterAid responded to COVID-19 in 26 countries implementing a do-no-harm phased response primarily focusing on hygiene behaviour change. The initial phase was based on non-contact methods such as promoting key hygiene behaviours through mass media, digital, social media and other non-contact methods. The response also included installation of handwashing facilities (mostly hands-free – paddle operated) with soap in public locations/institutions. In the second phase, communities were reached with non-contact face-to-face behaviour change activities (depending on in-country lockdown measures).

WaterAid developed a range of promotional package materials / assets / tools for the first phase of the campaign. In-person small group hygiene session assets and COVID-19 behaviours integrated into existing manuals were developed for community sessions which occurred, where possible, while maintaining physical distancing and wearing masks. As part of the first phase of the campaign, there were a broad range of promotional assets developed to expose target population multiple times with repeated frequency in all countries. There was a wide range to maintain interest; a celebrity presenting Emo-Demos of key behaviours on TV, mobilization of local FM and radios with key motivational messages, digital media using visual illustration of the behaviours to be performed, a series of radio dramas on local languages, and illustrations of key behaviours as picture/text or video clips shared via WhatsApp, Instagram, Facebook, Twitter, and TikTok. Additional initiatives include radio dramas, bill boards at travel entry/exit points, use of celebrity media slots, placement of visual cues or nudges next to handwashing facilities and print media in all the countries.

These promotional assets were developed in partnership with national governments and endorsed by government in many countries. For example, in Zambia, WaterAid worked with the Ministry of Health and other partners to facilitate the review of the new COVID-19 national intervention guidance package. In Nepal, WaterAid integrated COVID-19 preventive behaviours into the existing national hygiene through vaccination programme at national scale. WaterAid advised and supported the national behaviour change programme. Ghana, Ethiopia and Tanzania also worked with the a ‘creative team’ and contributed to developing and finalising the hygiene promotion materials.

WaterAid worked to support national governments to develop sustained behaviour-centred programmes under a national / regional / district programme. These programmes have their own identities such as a central theme, logo with government representation and authorisation. WaterAid’s hygiene response to COVID-19 was implemented within the following branded campaigns:

| **Ethiopia**  TSEDU  “Clean-Ethiopia” | **Ghana**  Clean community campaign | | **Zambia**  Kutuba (clean campaign) | | **Nigeria**  Clean Community Campaign (designed for Bauchhi State) |
| --- | --- | --- | --- | --- | --- |
|  | 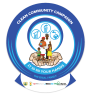 | | 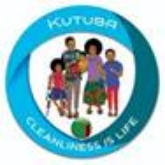 | | 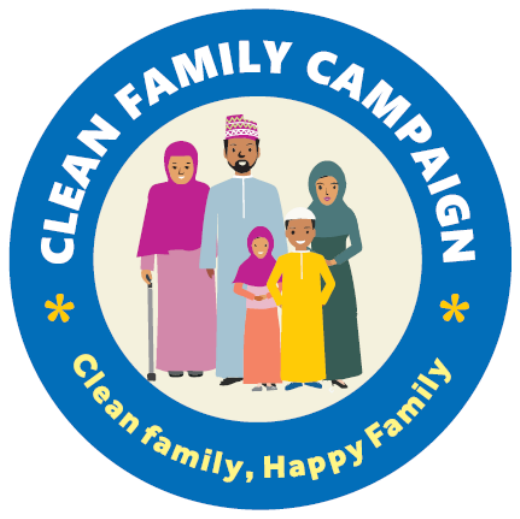 |
| **Tanzania**  National Behaviour Change Campaign on Hygiene and Sanitation  (Led by FCDO funded Clear Consortium with LSHTM) | | **Nepal**  Ideal family (clean family, happy family) campaign  (national hygiene into immunization) | | **Rwanda**  Shishoza Campaign  (branded specifically for COVID-19 response) | |
| 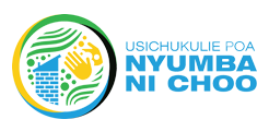 | | 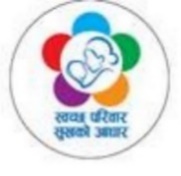 | | 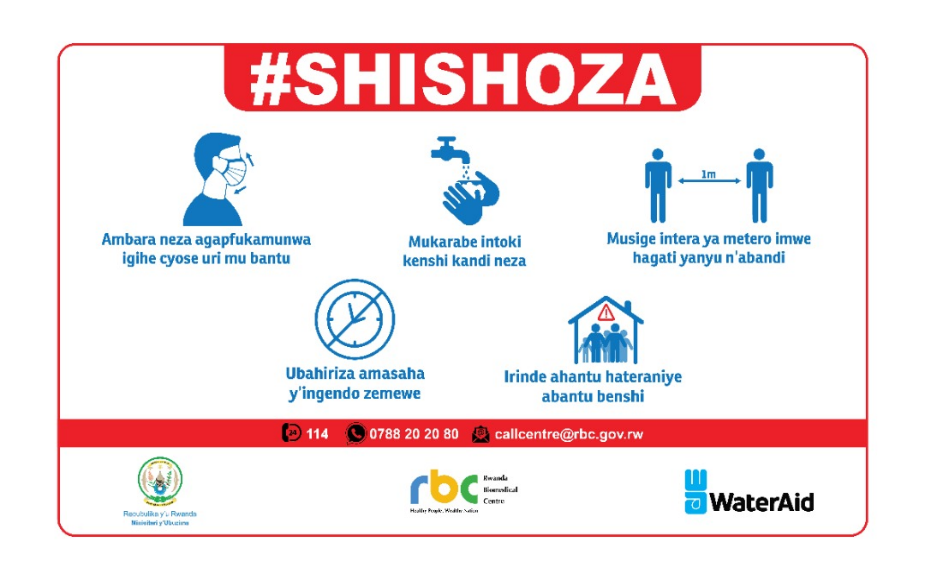 | |

#### Note S2: Sampling

For each country, the sampling frame for this survey is all adult (18+) members of households living within geographical areas supported by WaterAid in its COVID-19 response. The geographical areas defined are sub-national in nature. For a geographical area to be included in the study area WaterAid must have been delivering or planning to deliver its mass media campaign.

However, it was not logistically feasible in most countries to deliver a survey representative of all areas targeted by the mass media campaign. As such, some country programmes focussed on areas where community-based activities (including hygiene behaviour promotional activities, provision of handwashing infrastructure or hygiene products), were delivered, as opposed to all areas where people has been exposed to the mass media campaign.

In each country the sampling process differed slightly depending on resources, logistical constraints, population data availability and data requirements. In Ghana, Rwanda, Tanzania and Zambia either proportionate sampling or cluster sampling using probability proportionate to size to select the clusters was used to create a representative sample of the study area. In Ethiopia, Nepal and Nigeria, purposive sampling was applied for logistical reasons, meaning the results are not representative.

In all countries the minimum sample size was determined in accordance with Cochran’s formula, with a desired confidence level 95%, with expected frequency of 50%. As such, the minimum sample size was 384 from each country. However, in some countries additional resources were available allowing for an increased sample size.

Once the clusters at the lowest administrative level for which population data was readily available were selected, either a random walk was initiated (if the administrative unit was small enough) or units within the administrative unit were randomly selected for the random walk to be situated within.

To conduct the random walk the following methodology was used:

1. Randomly select a starting point in the community
   a. Go to some central location within the community (a market, a church, a health facility or the junction (if you have multiple enumerators in the same community drop them off in different locations)
   b. Select a direction at random by spinning a bottle. Move in a straight line in this direction.
   c. Generate a random number x (from pieces of paper numbered 1-10). Stop at the xth house as your first to form the starting point for the survey
2. Continue the random walk from your starting point
   a. Select a direction at random by spinning a bottle. Move in a straight line in this direction.
   b. Generate a random number x (from pieces of paper numbered 1-10). Stop at the xth house as your next household for the survey

In order to ensure an even gender balance in the sample, half of the enumerators were assigned to collect data from men, and half to collect data from women. On arriving at the household, the enumerator asked all adult respondents from the selected gender to draw straws to select the respondent to the questionnaire. If no one from the selected gender was present at the household, the random walk continued. If only one enumerator was present in the local area the gender of the respondent alternated each time.

1. Aunger R, Curtis V. *Behaviour Centred Design: towards an applied science of behaviour change*. Health Psychol Rev. 2016;10(4):425-46.
