## Supplementary File 2 for "Frequency and determinants of COVID-19 prevention behaviours: assessment of large-scale programmes in seven countries"

### Supplementary File 2: Questionnaires

**Introduction**

Hello! Good morning / afternoon / evening, my name is <Insert name and surname here> and I am from <insert name of organisation here>. We would like to invite you to take part in a survey. Before you decide if you want to participate, you need to understand why the survey is being done and what it would involve. I will go through information about the survey with you and answer any questions you may have. Let me know if anything I tell you is not clear or you would like more information.

**What is the purpose of this survey?**

As part of an ongoing response to COVID-19 and its associated hygiene campaign led by WaterAid in <insert name of district/region/country>, we are doing a survey to find out more information about COVID-19 and associated behaviours in this area in order to strengthen the programme and prevent the spread of COVID-19 and other diseases.

**Why have I been asked to take part?**

You have been invited because you live in one of the survey’s target areas.

**Do I have to take part?**

It is up to you to decide to take part or not. If you don’t want to take part, that’s ok. Your participation is voluntary. You can choose not to answer any question(s). And if you decide you do not want to continue, I will be happy to stop the interview at any point. For many of the questions that will be asked, there are no right or wrong answers – I want to know what you think and what are your practices.

**What will happen to me if I take part?**

For the survey, you will talk to us for about 45 - 60 minutes, after the consent process has been completed. As part of this process, I will ask you some socio-demographic information, and your understanding and exposure on key behaviours related to COVID-19 to prevent the spread of the virus and other diseases.

**What are the possible risks and disadvantages?**

You might find some of the questions we ask unusual or uncomfortable. If so, you do not have to answer these questions.

**What are the possible benefits?**

We cannot promise the survey will help you individually but the information we get from the survey will help our knowledge and understanding of coronavirus in your area.

**Can I change my mind about taking part?**

Yes. You can withdraw from the survey at any time. You are not obliged to participate in the survey and nothing will happen in consequence if you decide not to. Everything we discuss will be held in strict confidence.

**What will happen to information collected about me?**

All information collected about you will be kept private. Only the survey staff who check that the survey is being carried out properly will be allowed to look at information about you. Anonymised data will be shared with external third parties so that they can also learn from the results of this study. Anonymised data means that any information about you which leaves WaterAid, will not have your name or address on it so that you cannot be recognised or identified by this data.

**What will happen to the results of this survey?**

This survey is an operational assessment and all information provided thereby will be treated as confidential and not be utilised for any other purpose except strengthening the programme.

**Further information and contact details:**

If you would like any further information, please contact (provide name of enumerator or someone in charge of data collection) who can answer any questions you may have about the survey. You can call them on this number: (provide contact number). Thank you for taking time to listen to this information. If you think you will take part in the survey let us proceed to the consent taking.

**I confirm that I have understood the information read out to me for the survey named “Mid-term Rapid Assessment - Hygiene Behaviour Change in response to COVID-19”. I have had the opportunity to consider the information, ask questions and have these answered satisfactorily.**

1. **Yes**
2. **No**

**I understand that my consent is voluntary and that I am free to withdraw this consent at any time without giving any reason and without any consequence to me.**

1. **Yes**
2. **No**

**I understand that identifiable data about or from me will only be shared with the local WaterAid survey team and will be deleted following the study completion.**

1. **Yes**
2. **No**

**I understand that anonymised data about me may be shared with third party organisations, and that I will not be identifiable from this information**

1. **Yes**
2. **No**

**I agree to me taking part in the above named study**

1. **Yes**
2. **No**

**[For data collector] I confirm that I have read the information sheet dated <insert date and year> for the survey named 'Mid-term Rapid Assessment - Hygiene Behaviour Change in response to COVID-19'. I have given the participant an opportunity to consider the information and have answered any questions satisfactorily. I informed the participant their consent is voluntary and they are free to withdraw this consent at any time without giving any reason and without any consequence to them. I have informed the participant that data about/from participants may be shared directly with other researchers and the Ministry of Health and that the participants will not be identifiable from this information. I have informed the participant that data about them may be shared with third party organisations and that they will not be identifiable from this information. I confirm that the participant has consented to the above-named study.**

1. **Yes**
2. **No**

**Respondent ID:**

**Module A: Demographic**

| **Variable ID** | **Question** | **Responses** |
| --- | --- | --- |
| Socio-demographic information | | |
| A001 | Gender | 1. Male 2. Female 3. Transgender 4. Other/ Prefer not to say |
| A002 | How old are you? | ______ years |
| A003 | Country | Dropdown list: |
| A004 | Location  *(note to data collector: if you know the don’t need to ask if you are not sure, ask to confirm and circle the answer)* | 1. Urban 2. Urban (informal settlements) 3. Peri-Urban 4. Rural |
| A005 | I would like to ask you about the people that normally live in your household.  How many people live in your household? | Insert number |
| A006 | Can you tell me the total number of people in your household who are older than 60? | Insert number |
| A007 | How many people between 5 and 17 years old normally live in your household? | Insert number |
| A008 | How many children younger than 5 live in your households? | Insert number |
| A009 | Occupation of the respondent | 1. Professional/technical/managerial 2. Clerical 3. Sales and services 4. Skilled manual 5. Unskilled manual 6. Agriculture 7. Other specify: 8. Prefer not to say |
| A010 | What is your main household income source?  ***Note: choose only one major source of income*** | 1. Full-time employment 2. Part-time employment 3. Daily wages 4. Agricultural 5. Others (specify): 6. Prefer not to say |
| A011 | What is the average monthly spending on leisure activities and the average monthly saving (after paying all bills) of the household? | a) Insert number (leisure): ____________ b) Insert number (saving): _____________ c) Don’t know ­­­   *(once you collect this in local currently, pls translate this into US$)* |
| A012 | How many years of formal education did you attend? | 1. No formal education 2. Some primary school completed 3. Primary school completed 4. Some secondary school completed 5. Secondary school completed 6. Higher education completed |
| A013 | What is your ethnic group?  *Please consider local context. If question is seen as sensitive, then enumerator can skip.* | Specify: |
| A014 | What religion do you follow? | 1. Islam 2. Christianity 3. Hinduism 4. Buddhism 5. Other – please specify 6. Prefer not to say |
| A015 | Since the start of the COVID-19 pandemic has your household’s monthly income gone down, stayed the same, on increased? | 1. Gone down 2. Stayed the same 3. Increased |
| A016 | For the people in your household, which of the following have contributed to a decline in your monthly income | 1. Wages have been reduced 2. Work hours / days of work have decreased 3. Job has been lost / furloughed 4. Unable to work because of loss of childcare 5. Unable to work because of taking care of sick relative |
| A017 | Do you have access to and/or use the following? | 1. Internet 2. TV 3. Mobile phone/telephone 4. Radio 5. Social media (Facebook, Twitter, WhatsApp) 6. A computer 7. Electricity 8. Newspapers/Magazines etc. |
| **Access to basic services** | | |
| A018 | W1: What is the main source of drinking water for members of your household? | 1. Piped into dwelling 2. Piped into compound, yard or plot 3. Piped to neighbour 4. Public tap / standpipe 5. Tube well, borehole 6. Protected dug well 7. Unprotected dug well 8. Protected spring 9. Unprotected spring 10. Rainwater collection 11. Tanker-truck 12. Cart with small tank / drum 13. Water kiosk 14. Surface water (river, stream, dam, lake, pond, canal, irrigation channel) Bottled water 15. Sachet water 16. Other (please specify) |
|  | W2: What is the main source of water used by members of your household for other purposes, such as cooking and handwashing? | 1. Piped into dwelling 2. Piped into compound, yard or plot 3. Piped to neighbour 4. Public tap / standpipe 5. Tube well, borehole 6. Protected dug well 7. Unprotected dug well 8. Protected spring 9. Unprotected spring 10. Rainwater collection 11. Tanker-truck 12. Cart with small tank / drum 13. Water kiosk 14. Surface water (river, stream, dam, lake, pond, canal, irrigation channel) Bottled water 15. Sachet water 16. Other (please specify) |
| A019 | Where is the water collected from? | 1. In own dwelling 2. In own yard/plot 3. Elsewhere |
| A020 | How long time does it take to go there, get water and come back (including queuing at water point)? In minutes (00 if on site) | Insert number (minutes):____________ |
| A021 | Since the start of the COVID-19 pandemic has your access to water service gone down, stayed the same, increased? | 1. Gone down 2. Stayed the same 3. Increased |
| A022 | What kind of toilet facility do members of your household usually use? | 1. Flush or pour flush toilet that flush to a cistern, septic tank, pit or sewer system. 2. Pit latrine with slab 3. Flush or pour flush to open drain 4. Pit latrine without slab/open pit 5. Other (please specify): |
| A023 | Do you share this facility with others who are not members of your household? | 1. Yes 2. No |
| A024 | Where is this facility located? | 1. In own dwelling 2. In own yard/plot 3. Elsewhere |
| A025 | Since the start of the COVID-19 pandemic has your cleaning practice to toilet gone down, stayed the same, increased? | 1. Gone down 2. Stayed the same 3. Increased |
| **Disabilities** | | |
| A026 | Do you have any long-term illness, health problem or disability which limits your daily activities or the work you can do? | 1. Yes 2. No |
| A027 | Do you have difficulty seeing, even if wearing glasses? | 1. No - no difficulty 2. Yes – some difficulty 3. Yes – a lot of difficulty 4. Cannot do at all |
| A028 | Do you have difficulty hearing, even if using a hearing aid? | 1. No - no difficulty 2. Yes – some difficulty 3. Yes – a lot of difficulty 4. Cannot do at all |
| A029 | Do you have difficulty walking or climbing steps? | 1. No - no difficulty 2. Yes – some difficulty 3. Yes – a lot of difficulty 4. Cannot do at all |
| A030 | Do you have difficulty remembering or concentrating? | 1. No - no difficulty 2. Yes – some difficulty 3. Yes – a lot of difficulty 4. Cannot do at all |
| A031 | Do you have difficulty (with self-care such as) washing all over or dressing? | 1. No - no difficulty 2. Yes – some difficulty 3. Yes – a lot of difficulty 4. Cannot do at all |
| A032 | Using your usual (customary) language, do you have difficulty communicating, for example understanding or being understood? | 1. No - no difficulty 2. Yes – some difficulty 3. Yes – a lot of difficulty 4. Cannot do at all |
| A033 | Do any of your family member have any long-term illness, health problem or disability which limits their daily activities or the work they can do? | 1. Yes  2. No |

**Module B: Behaviours and Barriers**

| **Variable ID** | **Question** | **Responses** |
| --- | --- | --- |
| **Handwashing with soap** | | |
| Understanding / Knowledge | | |
| B040 | Think back to what you did yesterday. Can you please tell me all of the times that you washed your hands? | _______ times |
| B041 | You said approximately____ times. How many of these times did you wash your hands with soap? | ________ times |
| B042 | Where do members of your household most often wash their hands? | 1. Sink/tap in dwelling/yard/plot 2. Bucket with tap 3. Basin and/or jug 4. No specific place for handwashing in dwelling/yard/plot |
| B043 | What do you use to wash your hands? | 1. Water only 2. Bar soap 3. Liquid soap 4. Powder detergent 5. Soapy water 6. Ash 7. Alcohol based hand rub/sanitizer 8. Other |
| B044 | Right now, is there a place in your home where you could go to wash your hands? | 1. Yes 2. No |
| B045 | Is there water available at this location right now? | 1. Yes 2. No |
| B046 | Is there soap available at this location right now? | 1. Yes 2. No |
| B047 | What are the key moments when anyone should wash their hands with soap?  Do not read responses, but can prompt, any other times? | 1. Before eating 2. Before cooking 3. Before feeding a child 4. After cleaning a young child 5. After using the toilet 6. When entering or leaving the household or any other building 7. After coming into physical contact with anyone outside your household 8. After touching surfaces outside the home (e.g. door knobs, railing, money etc) 9. After sneezing or coughing 10. Before, during and after caring for a sick person Others – please specify |
| B048 | Any other information you can provide about how you should wash your hands?  (Multiple answers possible – tick all that are mentioned) | 1. For 20 seconds (or more)  2. Rubbing with both hands  3. Wash under running water  4. Save water  5. Avoid splashing  6. Avoid touching tap  7. Washing hands only with soap  8. Washing hands only with water  9. Other (please specify) |
| B049 | What are the benefits of washing hands with soap?  (note: multiple answer expected; just mention, what else?) | 1. Kills germs / prevent germs from spreading 2. Keep you healthy / avoid diseases 3. Keeps you clean / pure 4. Clean dust / dirt from hands 5. Makes you comfortable 6. Protect children 7. To be pure / clean / fresh 8. Prevents you from getting COVID-19 9. Others |
| Application / Current practice (reported) | | |
| B050 | On average, how long do you wash your hands for?  Record in seconds | Insert number (in seconds): |
| B051 | Do you have a way of counting to check the time while washing your hands? | 1. Yes – please describe  2. No |
| B052 | When do you wash your hands with soap and water?  *(Can have multiple answers. Don’t read but just mention what else?. Only circle the response if they use soap)* | 1. Before eating 2. Before cooking 3. Before feeding a child 4. After cleaning a young child 5. After using the toilet 6. When entering or leaving the household or any other building 7. After coming into physical contact with anyone outside your household 8. After touching surfaces outside the home (e.g. door knobs, railing, money etc) 9. After sneezing or coughing 10. Before, during and after caring for a sick person 11. Others – please specify |
| B053 | Since the outbreak of Covid-19 and as of now, has your handwashing with soap frequency per day decreased, stay the same or increased? | 1. Increased 2. Stayed the same 3. Decreased |
| B054 | Do you wash your hands outside the home? (at healthcare facilities, markets, ports of entry) | 1. Yes 2. No 3. Sometimes 4. Don’t know |
| B055 | Think about the last 3 times you went to a place outside the home. For example, at a healthcare facility, market or transport facility.  How many of these places had a place where you could wash your hands?? | Insert number |
| B056 | How many of these places had both soap and water available? | Insert number |
| B057 | Have you noticed a change in availability of handwashing facilities or soaps since Covid-19? | 1. Increased 2. Decreased 3. Stayed the same |
| Barriers | | |
| B058 | Think about the times in your day when you normally WANT to wash your hands. Are there things that sometimes make it difficult for you to wash your hands? | 1.Yes  2.No |
| B059 | I am going to ask you a series of questions about what people commonly tell us keeps them from washing hands as much as they know they should. Can you tell me if the following situations apply to you?  Soap that you use for handwashing is too expensive to purchase | 1.Yes  2.No |
| B060 | Soap that you use for handwashing is not available to purchase | 1. Yes  2. No |
| B061 | I need to save the soap I have for purposes other than handwashing | 1. Yes  2. No |
| B062 | Water is not available for handwashing | 1. Yes  2. No |
| B063 | Water is too expensive to purchase for handwashing | 1. Yes  2. No |
| B064 | The water I have is too dirty to use for handwashing | 1. Yes  2. No |
| B065 | I am too busy and do not have time to stop and wash my hands | 1. Yes  2. No |
| B066 | The place where I wash my hands is too far away | 1. Yes  2. No |
| B067 | I sometimes forget to wash my hands | 1. Yes  2. No |
| B068 | Program to auto select the ones that the participant has responded yes to – ask the participant to tell you which is the **most important** reason they do not always physically distance in public. |  |

| **Respiratory Hygiene (including wearing a mask)** | | |
| --- | --- | --- |
| Understanding / Knowledge | | |
| B069 | What do you think is meant by practicing ‘good respiratory hygiene’ linked to COVID19?  (Multiple answers possible – tick all that are mentioned) | 1. Covering nose and mouth when coughing/ sneezing  2. Sneezing or coughing into elbow  3. Wearing mask in public  4. Don’t know  5. Other – please specify |
| B070 | When should you wear a mask or face covering?  (Multiple answers possible – tick all that are mentioned) | 1. When in public places (markets, bus, train, healthcare centre, etc) 2. If I have possible Covid-19 symptoms 3. If I am taking care of someone who is ill 4. Don’t know 5. Other (please specify) |
| B071 | What is the usefulness of wearing mask in public?  (Multiple answers possible – tick all that are mentioned) | 1. To protect myself against Covid-19/ disease 2. To protect family and loved ones 3. To protect others while in public 4. Following instructions (e.g. from government, community or religious leaders) 5. To conform/ be respected by community or peers 6. Other – please specify |
| B072 | Do you know how to make a facemask? | 1. Yes  2. No  3. Don’t know |
| Application / current practice | | |
| B073 | Do you own a facemask or have something that you use to cover your mouth and nose in public? | 1. Yes  2. No  3. Don’t know |
| B074 | Do you wear a mask or face covering in public places? | 1. Always 2. Sometimes 3. Never |
| B075 | Do you cover your mouth when coughing or sneezing? | 1. Always 2. Sometimes 3. Never |
| B076 | Where do you dispose tissue if used after coughing or sneezing?  Note: Pick only one | 1. In closed bin with lid  2. In toilet  3. In waste collecting container  4. Through anywhere  5. Others; specify  6. Don’t use tissue yet all. |
| Barriers | | |
| B077 | Are there any barriers that prevent you from wearing a face mask in public? | 1. Yes 2. No |
| B078 | We would like to ask you about some of the potential things that may keep you from wearing a mask in public based on responses we have heard from other people. For each of these, please tell me if they apply to you:  I do not know where to find the materials to make a mask | 1. Yes  2. No |
| B079 | The material to make a mask is too expensive | 1. Yes  2. No |
| B080 | I do not know how to make a mask | 1. Yes  2. No |
| B081 | I do not know where I can purchase a mask | 1. Yes  2. No |
| B082 | The masks available for purchase are too expensive for me | 1. Yes  2. No |
| B083 | A face mask is too hot or too uncomfortable to wear | 1. Yes  2. No |
| B084 | I do not like the way I look when wearing a mask | 1. Yes  2. No |
| B085 | It is difficult to breathe when I am wearing a mask | 1. Yes  2. No |
| B086 | People will judge me if I am wearing a mask | 1. Yes  2. No |
| B087 | I simply forget my mask at home / do not have a mask available when I need it? | 1. Yes  2. No |
| B088 | Program to auto select the ones that the participant has responded yes to – ask the participant to tell you which is the **most important** reason they do not always wear a mask in public. |  |

| **Social / physical distancing** | | |
| --- | --- | --- |
| Understanding / Knowledge | | |
| B089 | What is meant by practicing physical / social distancing linked to COVID-19?  (*note: multiple answer expected - probe further, say what else. Don’t read the answer)* | 1. Staying at home 2. Staying 2 metres (or 1 metre – depends on country) from others 3. Not going to public places 4. Not socialising with other people 5. Don’t know |
| B090 | Why should we remain 2 metres (or 1 metre – depends on country) from others in public places and practice non-contact greetings?  (Multiple answers possible – tick all that are mentioned) | 1. To protect other people from getting Covid-19 2. To prevent us from getting Covid-19 3. It’s government rules, we have to follow 4. Don’t know 5. Other – please specify |
| Application / Current practices | | |
| B091 | When you leave your home – how often are you able to maintain physical distancing – which we define as staying at least (1 meter / 2 meters) away from other individuals  (Read responses) | 1. Always 2. Sometimes 3. Never 4. I don’t go out with my limitation |
| B092 | If your employer was required by the government to ensure employees physical distanced, do you think that would be possible at your workplace?  Note: only applicable when Option number 1 will be selected in A010)  (Note: Only relevant for people who said they were salaried?) | 1. Yes, easily 2. Yes, with some modifications 3. No 4. Other – please specify 5. Not applicable |
| Barriers | | |
| B093 | Are there times when it is difficult for you to maintain physical distancing? | 1. Yes  2. No |
| B094 | We would like to ask you about some of the potential things that may keep you from physically distancing in public based on responses we have heard from other people. For each of these, please tell me if they apply to you:  I don’t believe it is important to maintain physical distance | 1. Yes 2. No |
| B095 | Other people do not maintain their distance from me | 1. Yes 2. No |
| B096 | I have to spend time in queues where distancing is not feasible | 1. Yes 2. No |
| B097 | There are too many people / spaces are too crowded | 1. Yes 2. No |
| B098 | Other barriers to maintain distance from others. | Specify: __________________________ |
| B099 | Program to auto select the ones that the participant has responded yes to – ask the participant to tell you which is the **most important** reason they do not always physically distance in public. |  |

**Module C: Norms**

Instruct the participant to think about people in their community. You may want to adjust the reference group. In many cultures, including parts of South Asia (India, Nepal), community refers to the people of the same social group such as caste.

**Script: Think about 10 people in your community. I am going to ask you what you think these people commonly do (*pls note: remind them, they should think 10 people while answering in their own community*).**

| Social Norms | | |
| --- | --- | --- |
| C130 | How many of the people in your community always wash their hands with soap the appropriate time (critical times)? | 1. No body / none 2. Less than half of them 3. About half of them 4. Most of them 5. All of them |
| C131 | How many of these people do you think would wash their hands before entering a public space | 1. No body / none 2. Less than half of them 3. About half of them 4. Most of them 5. All of them |
| C132 | How many of them wear masks when they are in public spaces? | 1. No body / none 2. Less than half of them 3. About half of them 4. Most of them 5. All of them |
| C133 | How many of them cover their face when they cough or sneeze? | 1. No body / none 2. Less than half of them 3. About half of them 4. Most of them 5. All of them |
| C134 | How many of them maintain appropriate physical distancing (2m or 1m) when they are in public spaces? | 1. No body / none 2. Less than half of them 3. About half of them 4. Most of them 5. All of them |
| C135 | How many of them maintain appropriate physical distancing when they are socializing with other individuals? | 1. No body / none 2. Less than half of them 3. About half of them 4. Most of them 5. All of them |
| C136 | how many of them do you think would self-isolate if they are feeling unwell? | 1. No body / none 2. Less than half of them 3. About half of them 4. Most of them 5. All of them |
| C137 | How many of them do you think keep frequently touched surfaces in their home clean? | 1. No body / none 2. Less than half of them 3. About half of them 4. Most of them 5. All of them |
| C138 | How many of them do you think owns disinfectant? | 1. No body / none 2. Less than half of them 3. About half of them 4. Most of them 5. All of them |
| Think of three people who are the most important to you.  *Hint: Read out responses. For the following questions C139 – D172 you can tell the participant to think of their answers as a scale from 1 -5. Where 1 is not at all and 5 is extremely so.* | | |
| C139 | Would they approve if you always washed your hands with soap before eating? | 1. Not at all 2. Slightly 3. Moderately 4. Very 5. Extremely |
| C140 | How much do they approve if you always wash your hands with soap after coming home? | 1. Not at all 2. Slightly 3. Moderately 4. Very 5. Extremely |
| C141 | How much do they approve if you always wear a mask when going to public places? | 1. Not at all 2. Slightly 3. Moderately 4. Very 5. Extremely |
| C142 | How much do they approve if you always cover your mouth, when coughing or sneezing? | 1. Not at all 2. Slightly 3. Moderately 4. Very 5. Extremely |
| C143 | How much do they approve if you always keep at least 2m/1 meter distance from other people, when you are in public places? | 1. Not at all 2. Slightly 3. Moderately 4. Very 5. Extremely |
| C144 | How much do they approve if you disinfect frequently touched surfaces? | 1. Not at all 2. Slightly 3. Moderately 4. Very 5. Extremely |
| C145 | How much do they approve if you always stay at home, when you are unwell? | 1. Not at all 2. Slightly 3. Moderately 4. Very 5. Extremely |

**Module D: Motives**

Read responses for first five questions or until the respondent is answering spontaneously in response categories

| Motivations | | |
| --- | --- | --- |
| D150 | How afraid are you of contracting COVID-19?  (read the answer) | 1. Not at all 2. Slightly 3. Moderately 4. Very 5. Extremely |
| D151 | How much do you think that washing hands with soap protects you from contracting COVID-19? | 1. Not at all 2. Slightly 3. Moderately 4. Very 5. Extremely |
| D152 | How afraid are you that your loved ones contract COVID-19? | 1. Not at all 2. Slightly 3. Moderately 4. Very 5. Extremely |
| D153 | How much do you think that you protect your loved ones from COVID-19 when you wash hands with soap? | 1. Not at all 2. Slightly 3. Moderately 4. Very 5. Extremely |
| D154 | If you wash your hands with soap, how proud are you of yourself? | 1. Not at all 2. Slightly 3. Moderately 4. Very 5. Extremely |
| D155 | If you wash your hands with soap, how attractive do you feel to others? | 1. Not at all 2. Slightly 3. Moderately 4. Very 5. Extremely |
| D156 | If you wash your hands with soap, how clean do you feel to others? | 1. Not at all 2. Slightly 3. Moderately 4. Very 5. Extremely |
| D157 | What else would motivate you to practice handwashing with soap at critical times? | Specify: |
| D158 | How afraid are you that you might contract COVID-19 if someone next to you doesn’t wear a mask? | 1. Not at all 2. Slightly 3. Moderately 4. Very 5. Extremely |
| D159 | How much do you think your community looks up to you when you wear a mask? | 1. Not at all 2. Slightly 3. Moderately 4. Very 5. Extremely |
| D160 | How much do you think that you protect your loved ones from COVID-19 when you wear a mask in public places? | 1. Not at all 2. Slightly 3. Moderately 4. Very 5. Extremely |
| D161 | If you wear a mask are you proud of yourself? | 1. Not at all 2. Slightly 3. Moderately 4. Very 5. Extremely |
| D162 | What else would motivate you to wear mask in public? | 1. Specify: |
| D163 | Are you afraid that you or your family might contract COVID-19 if you do not clean/disinfect frequently touched surfaces? | 1. Not at all 2. Slightly 3. Moderately 4. Very 5. Extremely |
| D164 | How much do you think that you protect your loved ones from COVID-19 when you clean/disinfect frequently touched surfaces? | 1. Not at all 2. Slightly 3. Moderately 4. Very 5. Extremely |
| D165 | If you clean/disinfect frequently touched surfaces are you proud of yourself? | 1. Not at all 2. Slightly 3. Moderately 4. Very 5. Extremely |
| D166 | What else would motivate you to clean frequently touched surface? | 1. Specify: |
| D167 | How afraid are you that you might contract COVID-19 if someone next to you didn’t practice physical distancing (2m / 1m apart)? | 1. Not at all 2. Slightly 3. Moderately 4. Very 5. Extremely |
| D168 | How much do you think your community looks up to you when you maintain physical distancing? | 1. Not at all 2. Slightly 3. Moderately 4. Very 5. Extremely |
| D169 | How much do you think that you protect your loved ones and community members from COVID-19 when you maintain physical distancing? | 1. Not at all 2. Slightly 3. Moderately 4. Very 5. Extremely |
| D170 | If you maintain physical distancing while in public, are you proud of yourself as community appreciate this? | 1. Not at all 2. Slightly 3. Moderately 4. Very 5. Extremely |
| D171 | What else would motivate you to practice physical distancing in public. | 1. Specify: |
|  | Do you know about any guidelines set by the government? | 1. Yes 2. No |
| D172 | If Yes, how much would you say you follow guidelines set by your government? | 1. Not at all 2. Slightly 3. Moderately 4. Very 5. Extremely 6. Don’t know |

**Module E: Touchpoints**

| **Touchpoints / Messaging** | | |
| --- | --- | --- |
| E180 | Where do you usually get information?  Note: Pick only one | 1. Community/Religious leader 2. Health worker at health facility 3. Newspaper 4. Radio/FM 5. Television 6. Social Media 7. Loud speaker (Facebook, twitter, WhatsApp) 8. Word of mouth (family, friends, etc.) 9. NGO 10. Government 11. Other – specify |
| E181 | Where do you get the most trustworthy information about COVID-19 from?  Note: Pick only one | 1. Community/Religious leader 2. Health worker at health facility 3. Newspaper 4. Radio/FM 5. Television 6. Social Media (Facebook, twitter, WhatsApp) 7. Loud speaker 8. Word of mouth (family, friends, etc.) 9. NGO 10. Government 11. Other – specify |
| E182 | Have you seen or heard any messages about behaviours or things you can do to protect yourself and limited the spread of COVID-19? | 1. Yes 2. No |
| E183 | If yes, what were the messages you have heard?  (Multiple answers possible – tick all that are mentioned) | 1. Handwashing hands with soap 2. Wearing mask in public 3. Maintaining physical distancing between people (1 to 2m) 4. Cleaning frequently touched surface 5. Coughing / sneezing in elbow 6. Staying at home when feeling unwell.  2. Visiting local health institutions when feeling unwell. 3. Symptoms of COVID-19 4. I don’t know 5. Seeking support from traditional healer |
| E184 | If yes, where did you see or hear this message?  Note for interviewer  Don’t read out list of answers (but if they only mention one source, you can prompt “any others”?)  (Multiple answers possible – tick all that are mentioned) | 1. Community/Religious leader 2. Health worker at health facility 3. Newspaper 4. Radio/FM 5. Television 6. Social Media 7. Loud speaker 8. Word of mouth (family, friends, etc.) 9. NGO 10. Visual promotional material (poster, pamphlet, etc) 11. Other specify:_____________ |
| E185 | Over the last two weeks, how often were you exposed to these messages? | 1. Once 2. 2-5 times 3. More than 5 times |
| E186 | What did you recall from the message?  (Multiple answers possible) | 1. The signs and symptoms of Covid-19 2. The behaviours that I should do to protect myself and others from getting Covid-19 3. The emotions that the advert made me feel 4. Characters and the story line from the advert 5. Don’t remember |
| E187 | Was there any message or anything which you did not understand? | 1. Yes 2. No |
| E188 | If yes why?  (Multiple answers possible – tick all that are mentioned) | 1. Too many different messages 2. Message/picture not clear or easily understood 3. Too busy to read/listen 4. Didn’t seem relevant 5. Unable to read easily (too small, unclear writing) 6. Unable to read (blind or partially sighted) 7. Unable to read – wrong language 8. Unable to read – illiterate 9. Unable to hear properly 10. Unable to hear – wrong language 11. Not sure 12. Other – please specify |
| E189 | Have you changed your handwashing behaviour as a result of these messages/advert? | 1. Yes 2. No 3. Don’t know 4. Others…… |
| E190 | If YES, what caused you to change?  (Multiple answers possible – tick all that are mentioned) | 1. I want to protect myself against Covid-19/disease 2. To protect family and loved ones 3. Desire for cleanliness 4. Following instructions (e.g. from government, community or religious leaders) 5. To conform/be respected by community or peers 6. Increased availability of handwashing facilities 7. Other – please specify |
| E191 | As a result of these messages/promotional, would you say you have made any changes to your respiratory hygiene practice or wearing a mask (or face cloth) in institutions (eg health-care facilities)? | 1. Yes, fully 2. Yes, somewhat 3. No 4. Don’t know |
| E192 | Have you heard messages / promotional content about social / physical distancing? | 1. Yes  2. No |
| E193 | If yes, as a result of these messages/promotion, would you say you have made any changes to your behaviours when in public places (market places, queuing for water, etc) | 1. Yes  2. No |
| E194 | Have you heard messages about staying at home if you feel unwell? | 1. Yes  2. No |
| E195 | If yes, as a result of these messages, would you say you personally have made any changes to your behaviours when it comes to self-isolating? | 1. Yes  2. No |
| E196 | Have you heard messages / promotion about cleaning and disinfecting frequently touched surfaces regularly such as door handle, mobile phone, light switches? | 1. Yes  2. No |
| E197 | As a result of these messages, would you say you started to clean frequently touched surfaces? | 1. Yes  2. No |
| E198 | Have you heard or seen any messages / advert / illustration from WaterAid about COVID19 and associated behaviours such as handwashing with soap, wearing a face masks, physical and social distancing, self-isolation or surface cleaning?  *(pls mention specific campaign name if any or government campaign if designed and name accordingly – if there is a specific famous character used you also further probe to remind)* | 1. Yes  2. No |
| E199 | If yes, what was the message(s) from WaterAid about?  Tick all that apply | 1. Handwashing with soap 2. Wearing a face mask 3. Physical and social distancing 4. Sneezing / coughing in elbow or using tissue and disposing used tissue in closed bin. 5. Self-isolation 6. Surface cleaning 7. About COVID19 symptoms 8. Other specify____________ 9. None of the above |
| E200 | Over the last two weeks, how often were you exposed to these messages? | 1. Once 2. 2-5 times 3. More than 5 times |
| E201 | If yes, where did you see/hear this message?  (Multiple answers possible – tick all that are mentioned) You can prompt anywhere else to check if all have been mentioned | 1. Community/Religious leader 2. Health worker at health facility 3. Newspaper 4. Radio/FM 5. Television 6. Social Media 7. Loud Speaker 8. Word of mouth (family, friends, etc.) 9. NGO 10. Government 11. Other – specify |
| E202 | In your preference, what is the best way to receive important messages / information about COVID19 / health issues?  Note: Pick only one | 1. Community/Religious leader 2. Health worker at health facility 3. Newspaper 4. Radio/FM 5. Television 6. Social Media 7. Loud speaker 8. Word of mouth (family, friends, etc.) 9. NGO 10. Government 11. Other specify:_________________ |

**Module F: COVID-19 Questions**

| COVID-19 | | |
| --- | --- | --- |
| F210 | Have you had one or more of the COVID-19 symptoms in the last 6 months? (shortness of breath, loss of taste, fever, new and continuous cough) | 1. Yes 2. No 3. Prefer not to say |
| F211 | Did you get tested for COVID-19? | 1. Yes 2. No 3. Prefer not to say |
| F212 | If now, why did you not get tested | 1. Wasn’t eligible for a test 2. The test was too expensive 3. I didn’t know where to get a test 4. I was self-isolating 5. Other specify: ___ |
| F213 | If yes, have you tested positive for COVID-19? | 1. Yes 2. No 3. Don’t know 4. Prefer not to say |
| F214 | Has anyone in your household tested positive for COVID-19? | 1. Yes 2. No 3. Don’t know 4. Prefer not to say |
| F215 | Do you know how COVID-19 can be prevented or how to reduce spreading? | 1. Yes 2. No 3. Don’t know |
| F216 | If yes, tell us what preventive measures can be done to protect yourself?  (Note: multiple answers are expected. Probe further) | 1. Handwashing hands with soap 2. Wearing mask in public 3. Maintaining physical distancing between people (1 to 2m) 4. Cleaning frequently touched surface 5. Coughing / sneezing in elbow 6. Staying at home when feeling unwell. 7. Visiting local health institutions when feeling unwell. 8. I don’t know 9. Seeking support from traditional healer 10. Others ----------------------------- |
| F217 | If yes, tell us what preventative measures can be done to prevent the spread to others?  (Note: multiple answers are expected. Probe further) | 1. Handwashing hands with soap 2. Wearing mask in public 3. Maintaining physical distancing between people (1 to 2m) 4. Cleaning frequently touched surface 5. Coughing / sneezing in elbow 6. Staying at home when feeling unwell. 7. Visiting local health institutions when feeling unwell. 8. I don’t know 9. Seeking support from traditional healer 10. Others ----------------------------- |

**Closure**

That completes the survey, thank you very much for your help.

After finishing the interview, give the following messages to the respondents

**COVID-19 Preventive Measure**

- Frequently wash your hands with soap and water for at least 20 sec
- Cover your mouth and nose with tissue or with bent when you cough or sneeze
- Maintain physical distancing at least two meter (six feet) (1m where recommended) from each other while in public
- Wear mask in public. If you are also supporting people with suspected symptoms
- Avoid touching face (mouth, nose and eyes) frequently.
- Avoid spitting in public
- Avoid mass gatherings
- Stay at home or seek immediate medical support when you have symptoms

**For Any Suspected Cases**

- High Fever, new Cough, loss of taste/smell or breathing difficulties
- Coming from COVID-19 affected countries
- Coming in contact with any COVID-19 positive cases

**For more information about COVID19**

PLEASE CALL -Corona Info Hotline: Fill in with information from specific country
