## Supplementary File 3 for "Frequency and determinants of COVID-19 prevention behaviours: assessment of large-scale programmes in seven countries"

### Supplementary File 3: Supplementary tables

#### Table S1: Construction of primary outcomes and determinants for the key behaviours

| **Construct** | **Items** | **PCA eigenvalue** | **Cronbach's ⍺** | **Final outcome** |
| --- | --- | --- | --- | --- |
| **GENERAL** |  |  |  |  |
| ***Motives*** | | | | |
| Fear of COVID-19 | How afraid are you of contracting COVID-19? |  | 0.84 | Index 0–3 |
|  | How afraid are you that your loved ones contract COVID-19? |  |  |  |
| **HWWS** |  |  |  |  |
| ***Primary outcomes*** | | | | |
| HWWS at key moments | *The following items are reported as key moments at which the respondent practices HWWS:* |  |  |  |
| COVID-19 HWWS index | When entering or leaving the household or any other building | 3.29 | 0.73 | Index 0–3 |
|  | After coming into physical contact with anyone outside your household |  |  |  |
|  | After touching surfaces outside the home (e.g. door knobs, railing, money etc) |  |  |  |
|  | After sneezing or coughing |  |  |  |
|  | Before, during and after caring for a sick person |  |  |  |
| After toilet use | After using the toilet |  | | Binary |
| Before eating | Before eating |  | | Binary |
| Increase in HWWS | Since the outbreak of Covid-19 and as of now, reported HWWS frequency per day has increased |  | | Binary |
| ***Knowledge*** | | | | |
| Action knowledge for HWWS | Washing hands with soap reported as a measure that can be done to protect self from COVID-19 |  | | Binary |
| Procedural knowledge for HWWS | *The following items are reported as key moments when anyone should practice HWWS:* | | | |
| COVID-19 prevention | When entering or leaving the household or any other building | 3.25 | 0.72 | Index 0–3 |
|  | After coming into physical contact with anyone outside your household |  |  |  |
|  | After touching surfaces outside the home (e.g. door knobs, railing, money etc) |  |  |  |
|  | After sneezing or coughing |  |  |  |
|  | Before, during and after caring for a sick person |  |  |  |
| After toilet use | After using the toilet |  | | Binary |
| Before eating | Before eating |  | | Binary |
| ***Barriers*** | | | | |
| Soap | Soap that you use for handwashing is too expensive to purchase | 2.39 | 0.67 | Index 0–3 |
|  | Soap that you use for handwashing is not available to purchase |  |  |  |
|  | I need to save the soap I have for purposes other than handwashing |  |  |  |
| Water | Water is not available for handwashing | 2.52 | 0.72 | Index 0–3 |
|  | Water is too expensive to purchase for handwashing |  |  |  |
|  | The water I have is too dirty to use for handwashing |  |  |  |
| Self-regulation | I am too busy and do not have time to stop and wash my hands | 2.18 | 0.52 | Index 0–3 |
|  | The place where I wash my hands is too far away |  |  |  |
|  | I sometimes forget to wash my hands |  |  |  |
| ***Norms*** | | | | |
| Descriptive norm | Think about 10 people in your community. How many of the people in your community always wash their hands with soap the appropriate time (critical times)? |  | 0.83 | Index 0–3 |
|  | Think about 10 people in your community. How many of these people do you think would wash their hands before entering a public space? |  |  |  |
| Injunctive norm | Think of three people who are the most important to you. Would they approve if you always washed your hands with soap before eating? |  | 0.85 | Index 0–3 |
|  | Think of three people who are the most important to you. How much do they approve if you always wash your hands with soap after coming home? |  |  |  |
| ***Motives*** | | | | |
| Belief that HWWS protects others from COVID-19 | How much do you think that washing hands with soap protects you from contracting COVID-19? |  | 0.85 | Index 0–3 |
|  | How much do you think that you protect your loved ones from COVID-19 when you wash hands with soap? |  |  |  |
| Pride in practicing HWWS | If you wash your hands with soap, how proud are you of yourself? |  | | Index 0–3 |
| Belief that HWWS makes respondent attractive to others | If you wash your hands with soap, how attractive do you feel to others? |  | | Index 0–3 |
| Belief that HWWS makes respondent clean to others | If you wash your hands with soap, how clean do you feel to others? |  | | Index 0–3 |
| **MASK WEARING** |  |  |  |  |
| ***Primary outcomes*** | | | | |
| Always wears a mask | Respondent reports always wearing a mask or face covering in public places |  | | Binary |
| ***Knowledge*** | | | | |
| Action knowledge for mask wearing | Wearing a mask in public reported as a measure that can be done to protect self from COVID-19 |  | | Binary |
| Procedural knowledge for mask wearing | *The following reported as situations where one should wear a mask or covering:* | | | |
|  | When in public places (markets, bus, train, healthcare centre, etc) | 1.73 | 0.44 | Index 0–3 |
|  | If I have possible Covid-19 symptoms |  |  |  |
|  | If I am taking care of someone who is ill |  |  |  |
| ***Barriers*** | | | | |
| Availability | I do not know where to find the materials to make a mask | 3.13 | 0.67 | Index 0–3 |
|  | The material to make a mask is too expensive |  |  |  |
|  | I do not know how to make a mask |  |  |  |
|  | I do not know where I can purchase a mask |  |  |  |
|  | The masks available for purchase are too expensive for me |  |  |  |
| Comfort | A face mask is too hot or too uncomfortable to wear | 2.33 | 0.69 | Index 0–3 |
|  | I do not like the way I look when wearing a mask |  |  |  |
|  | It is difficult to breathe when I am wearing a mask |  |  |  |
| Pride | People will judge me if I am wearing a mask |  | | Binary |
| Self-regulation | I simply forget my mask at home / do not have a mask available when I need it |  | | Binary |
| ***Norms*** | | | | |
| Descriptive norm | Think about 10 people in your community. How many of them wear masks when they are in public spaces? |  | | Index 0–3 |
| Injunctive norm | Think of three people who are the most important to you. How much do they approve if you always wear a mask when going to public places? |  | | Index 0–3 |
| ***Motives*** | | | | |
| Fear of contracting COVID-19 if mask wearing is not practiced | How afraid are you that you might contract COVID-19 if someone next to you doesn’t wear a mask? |  | | Index 0–3 |
| Belief that mask wearing protects others from COVID-19 | How much do you think that you protect your loved ones from COVID-19 when you wear a mask in public places? |  | | Index 0–3 |
| Pride in practicing mask wearing | If you wear a mask are you proud of yourself? |  | | Index 0–3 |
| Respect from community for practicing mask wearing | How much do you think your community looks up to you when you wear a mask? |  | | Index 0–3 |
| **PHYSICAL DISTANCING** |  |  |  |  |
| ***Primary outcomes*** | | | | |
| Always physical distancing | Respondent reports always being able to maintain physical distancing – staying at least (1 meter / 2 meters) away from other individuals – when leaving the home |  | | Binary |
| ***Knowledge*** | | | | |
| Action knowledge for physical distancing | Maintaining physical distancing between people (1 to 2m) reported as a measure that can be done to protect self from COVID-19 |  | | Binary |
| Procedural knowledge for physical distancing | Staying 2 metres (or 1 metre – depends on country) from others reported as meaning of physical distancing |  | | Binary |
| ***Barriers*** | | | | |
| Response efficacy | I don’t believe it is important to maintain physical distance |  | | Binary |
| Space | Other people do not maintain their distance from me | 2.41 | 0.73 | Index 0–3 |
|  | I have to spend time in queues where distancing is not feasible |  |  |  |
|  | There are too many people / spaces are too crowded |  |  |  |
| ***Norms*** | | | | |
| Descriptive norm | Think about 10 people in your community. How many of them maintain appropriate physical distancing (2m or 1m) when they are in public spaces? |  | 0.87 | Index 0–3 |
|  | Think about 10 people in your community. How many of them maintain appropriate physical distancing when they are socializing with other individuals? |  |  |  |
| Injunctive norm | Think of three people who are the most important to you. How much do they approve if you always keep at least 2m/1 meter distance from other people, when you are in public places? |  | | Index 0–3 |
| ***Motives*** | | | | |
| Fear of contracting COVID-19 if physical distancing is not practiced | How afraid are you that you might contract COVID-19 if someone next to you didn’t practice physical distancing (2m / 1m apart)? |  | | Index 0–3 |
| Belief that physical distancing protects others from COVID-19 | How much do you think that you protect your loved ones and community members from COVID-19 when you maintain physical distancing? |  | | Index 0–3 |
| Pride in practicing physical distancing | If you maintain physical distancing while in public, are you proud of yourself as the community appreciates this? |  | | Index 0–3 |
| Respect from community for practicing physical distancing | How much do you think your community looks up to you when you maintain physical distancing? |  | | Index 0–3 |

#### Table S2: Demographic characteristics of respondents and their households by country

| **Country** | **GLOBAL** | **Ethiopia** | **Ghana** | **Nepal** | **Nigeria** | **Rwanda** | **Tanzania** | **Zambia** |
| --- | --- | --- | --- | --- | --- | --- | --- | --- |
| **Individuals** | 3033 | 505 | 387 | 497 | 422 | 423 | 395 | 404 |
| **Villages** | 211 | 8 | 39 | 25 | 48 | 47 | 11 | 33 |
| **Geographic area** |  |  |  |  |  |  |  |  |
| Urban | 1302 (42.9) | 505 (100) | 0 (0.0) | 144 (29.0) | 183 (43.4) | 85 (20.1) | 154 (39.0) | 231 (57.2) |
| Peri-Urban | 712 (23.5) | 0 (0.0) | 90 (23.3) | 160 (32.2) | 149 (35.3) | 106 (25.1) | 154 (39.0) | 53 (13.1) |
| Rural | 1019 (33.6) | 0 (0.0) | 297 (76.7) | 193 (38.8) | 90 (21.3) | 232 (54.9) | 87 (22.0) | 120 (29.7) |
| **Gender** |  |  |  |  |  |  |  |  |
| Male | 1469 (48.4) | 250 (49.5) | 193 (49.9) | 253 (50.9) | 253 (60.0) | 189 (44.7) | 160 (40.5) | 171 (42.3) |
| Female | 1564 (51.6) | 255 (50.5) | 194 (50.1) | 244 (49.1) | 169 (40.1) | 234 (55.3) | 235 (59.5) | 233 (57.7) |
| **Age** |  |  |  |  |  |  |  |  |
| 15-25 | 442 (14.6) | 71 (14.1) | 81 (20.9) | 65 (13.1) | 33 (7.9) | 34 (8.1) | 62 (15.7) | 96 (23.8) |
| 26-50 | 1964 (65.0) | 328 (65.1) | 199 (51.4) | 327 (65.9) | 293 (69.9) | 292 (69.5) | 256 (65.0) | 269 (66.8) |
| >50 | 617 (20.4) | 105 (20.8) | 107 (27.7) | 104 (21.0) | 93 (22.2) | 94 (22.4) | 76 (19.3) | 38 (9.4) |
| *Missing (impossible values)* | *10 (0.3)* | *1 (0.2)* | *0 (0.0)* | *1 (0.2)* | *3 (0.7)* | *3 (0.7)* | *1 (0.3)* | *1 (0.3)* |
| **Highest education level** |  |  |  |  |  |  |  |  |
| Primary not completed | 962 (31.9) | 147 (29.3) | 198 (51.2) | 218 (43.9) | 65 (15.4) | 203 (48.5) | 74 (18.9) | 57 (14.2) |
| Primary school completed | 971 (32.2) | 123 (24.6) | 101 (26.1) | 156 (31.4) | 110 (26.1) | 179 (42.7) | 198 (50.5) | 104 (25.9) |
| Secondary school or higher completed | 1086 (36.0) | 231 (46.1) | 88 (22.7) | 123 (24.8) | 246 (58.4) | 37 (8.8) | 120 (30.6) | 241 (60.0) |
| *Missing* | *14 (0.5)* | *4 (0.8)* | *0 (0.0)* | *0 (0.0)* | *1 (0.2)* | *4 (1.0)* | *3 (0.8)* | *2 (0.5)* |
| **Respondent has long term illness or disability** | 745 (24.6) | 129 (25.5) | 110 (28.4) | 150 (30.2) | 68 (16.1) | 122 (28.8) | 94 (23.8) | 72 (17.8) |
| **Family member has long term illness or disability** | 701 (23.2) | 114 (22.9) | 88 (22.7) | 165 (33.2) | 56 (13.3) | 72 (17.1) | 71 (18.1) | 135 (33.4) |
| *Missing* | *15 (0.5)* | *8 (1.6)* | *0 (0.0)* | *0 (0.0)* | *1 (0.2)* | *3 (0.7)* | *3 (0.8)* | *0 (0.0)* |
| **Household has children under 5** | 1555 (53.0) | 172 (34.2) | 264 (68.2) | 187 (39.7) | 286 (76.1) | 222 (53.4) | 220 (56.4) | 204 (52.2) |
| *Missing* | *99 (3.3)* | *2 (0.4)* | *0 (0.0)* | *26 (5.2)* | *46 (10.9)* | *7 (1.7)* | *5 (1.3)* | *13 (3.2)* |
| **Household has members over 60** | 1053 (36.0) | 154 (30.8) | 225 (58.4) | 213 (44.5) | 164 (47.1) | 76 (18.1) | 118 (30.0) | 103 (25.8) |
| *Missing* | *106 (3.5)* | *5 (1.0)* | *2 (0.5)* | *18 (3.6)* | *74 (17.5)* | *2 (0.5)* | *1 (0.3)* | *4 (1.0)* |

#### Table S3: Household access to water, sanitation and hygiene services by country

| **Country** | **GLOBAL** | **Ethiopia** | **Ghana** | **Nepal** | **Nigeria** | **Rwanda** | **Tanzania** | **Zambia** |
| --- | --- | --- | --- | --- | --- | --- | --- | --- |
| **N** | 3033 | 505 | 387 | 497 | 422 | 423 | 395 | 404 |
| **Access to drinking water** | | |  |  |  |  |  |  |
| Safely managed | 1694 (57.7) | 441 (87.9) | 75 (20.5) | 429 (86.5) | 168 (45.4) | 108 (25.8) | 220 (57.6) | 253 (62.8) |
| Basic | 643 (21.9) | 24 (4.8) | 187 (51.1) | 51 (10.3) | 124 (33.5) | 69 (16.5) | 85 (22.3) | 103 (25.6) |
| Limited | 310 (10.6) | 30 (6.0) | 61 (16.7) | 4 (0.8) | 20 (5.4) | 104 (24.9) | 64 (16.8) | 27 (6.7) |
| Unimproved | 165 (5.6) | 2 (0.4) | 6 (1.6) | 12 (2.4) | 52 (14.1) | 70 (16.8) | 13 (3.4) | 10 (2.5) |
| No service | 125 (4.3) | 5 (1.0) | 37 (10.1) | 0 (0.0) | 6 (1.6) | 67 (16.0) | 0 (0.0) | 10 (2.5) |
| *Missing* | *96 (3.2)* | *3 (0.6)* | *21 (5.4)* | *1 (0.2)* | *52 (12.3)* | *5 (1.2)* | *13 (3.3)* | *1 (0.3)* |
| **Access to sanitation** | |  |  |  |  |  |  |  |
| At least basic | 1799 (59.7) | 279 (55.7) | 94 (24.3) | 401 (80.7) | 245 (58.8) | 345 (82.3) | 197 (50.8) | 238 (59.1) |
| Limited | 618 (20.5) | 145 (28.9) | 64 (16.5) | 59 (11.9) | 92 (22.1) | 36 (8.6) | 145 (37.4) | 77 (19.1) |
| Unimproved | 379 (12.6) | 77 (15.4) | 51 (13.2) | 34 (6.8) | 48 (11.5) | 37 (8.8) | 46 (11.9) | 86 (21.3) |
| No service | 216 (7.2) | 0 (0.0) | 178 (46.0) | 3 (0.6) | 32 (7.7) | 1 (0.2) | 0 (0.0) | 2 (0.5) |
| *Missing* | *21 (0.7)* | *4 (0.8)* | *0 (0.0)* | *0 (0.0)* | *5 (1.2)* | *4 (1.0)* | *7 (1.8)* | *1 (0.3)* |
| **Access to handwashing facilities** | |  |  |  |  |  |  |  |
| Basic (soap and water) | 1498 (50.0) | 235 (47.7) | 138 (35.8) | 422 (84.9) | 165 (39.9) | 210 (50.6) | 111 (28.5) | 217 (54.3) |
| Limited | 258 (8.6) | 47 (9.5) | 9 (2.3) | 55 (11.1) | 24 (5.8) | 24 (5.8) | 64 (16.4) | 35 (8.8) |
| No service | 1239 (41.4) | 211 (42.8) | 239 (61.9) | 20 (4.0) | 225 (54.4) | 181 (43.6) | 215 (55.1) | 148 (37.0) |
| *Missing* | *38 (1.3)* | *12 (2.4)* | *1 (0.3)* | *0 (0.0)* | *8 (1.9)* | *8 (1.9)* | *5 (1.3)* | *4 (1.0)* |

#### Table S4: Global prevalence of primary outcomes disaggregated by key demographics

|  | **Moments for COVID-19 prevention (0-3): mean (SD)** | **After toilet use: n (%)** | **Before eating: n (%)** | **Increased after COVID-19:**  **n (%)** | **Always wears mask in public spaces: n (%)** | **Always physically distances in public spaces: n (%)** |
| --- | --- | --- | --- | --- | --- | --- |
| **N** | **3033** | | | | | |
| **Outcome at global level** | 0.96 (1.06) | 2518 (83.3) | 2802 (92.7) | 2415 (80.2) | 1757 (58.3) | 864 (29.4) |
| ***Disaggregated by:*** |  |  |  |  |  |  |
| **Geographic area** |  |  |  |  |  |  |
| Urban | 0.98 (1.04) | 1021 (78.6) | 1202 (92.5) | 1039 (80.7) | 678 (52.6) | 308 (24.1) |
| Peri-Urban | 0.79 (0.98) | 598 (84.1) | 657 (92.4) | 528 (74.4) | 415 (58.5) | 160 (23.5) |
| Rural | 1.04 (1.12) | 899 (88.7) | 943 (93.0) | 848 (83.5) | 664 (65.4) | 396 (40.3) |
| **Gender** |  |  |  |  |  |  |
| Male | 1.04 (1.08) | 1215 (83.0) | 1375 (94.0) | 1156 (79.3) | 839 (57.5) | 393 (27.3) |
| Female | 0.88 (1.03) | 1303 (83.5) | 1427 (91.4) | 1259 (80.9) | 918 (59.0) | 471 (31.4) |
| **Age group** |  |  |  |  |  |  |
| 15-25 | 0.85 (1.02) | 365 (83.0) | 401 (91.1) | 338 (76.8) | 245 (55.8) | 105 (24.4) |
| 26-50 | 0.95 (1.05) | 1645 (83.9) | 1813 (92.5) | 1590 (81.4) | 1117 (57.2) | 563 (29.5) |
| > 50 | 1.04 (1.09) | 499 (81.4) | 578 (94.3) | 479 (78.5) | 389 (63.5) | 192 (32.6) |
| **Respondent has long term illness or disability** | |  |  |  |  |  |
| No | 0.93 (1.05) | 1932 (84.7) | 2123 (93.1) | 1859 (81.8) | 1299 (57.1) | 627 (28.1) |
| Yes | 1.02 (1.08) | 586 (78.8) | 679 (91.3) | 556 (75.0) | 458 (61.8) | 237 (33.4) |
| **Household relative wealth quintile** | |  |  |  |  |  |
| Lowest | 0.94 (1.11) | 530 (74.2) | 654 (91.6) | 488 (68.6) | 327 (46.1) | 172 (25.1) |
| Second | 0.86 (1.00) | 463 (81.8) | 518 (91.5) | 470 (82.6) | 325 (57.4) | 156 (28.6) |
| Middle | 0.97 (1.06) | 542 (87.3) | 574 (92.4) | 516 (83.6) | 347 (56.1) | 190 (31.4) |
| Fourth | 0.89 (0.98) | 499 (86.9) | 546 (95.1) | 478 (84.2) | 348 (60.8) | 151 (26.6) |
| Highest | 1.15 (1.10) | 452 (89.7) | 472 (93.7) | 424 (84.3) | 376 (74.6) | 176 (35.6) |
